# Time to diagnosis among children and adolescents with cancer in Quebec, Canada: a population-based study

**DOI:** 10.64898/2026.04.09.26350491

**Authors:** Callum Mullen, Ronald D. Barr, Erin Strumpf, Mariam El-Zein, Eduardo L. Franco, Talía Malagón

**Affiliations:** Gerald Bronfman Department of Oncology, Division of Cancer Epidemiology, McGill University, Montreal, QC, Canada; Department of Epidemiology, Biostatistics and Occupational Health, McGill University, Montreal, QC, Canada; Department of Pediatrics, McMaster University, Hamilton, ON, Canada; Department of Economics, McGill University, Montreal, QC, Canada; St Mary’s Research Centre, Montréal West Island CIUSSS, Montreal, QC, Canada

**Author notes:** **Corresponding author:** Talía Malagón.

## Abstract

**Background:** Timely cancer diagnosis in children and adolescents is critical to improving outcomes, yet substantial variation in diagnostic intervals persists across cancer types and care settings. We aimed to quantify time to diagnosis and assess variations by patient, demographic, and system-level factors.

**Methods:** We conducted a retrospective population-based study of children and adolescents aged 0–19 years diagnosed with one of 12 common cancers between 2010 and 2022 in Quebec, Canada. The diagnostic interval was defined as the time from first cancer-related healthcare encounter to diagnosis. We calculated medians and interquartile ranges (IQR) overall and by cancer type and used multivariable quantile regression to identify factors associated with time to diagnosis at the 25^th^, 50^th^, and 75^th^ percentiles.

**Results:** Among 2,927 individuals with cancer, diagnostic intervals varied by cancer type and age. Median intervals were longest for carcinomas (100 days; IQR 33–192) and shortest for leukemias (8 days; IQR 3–44). Compared with children living in Montréal, living in regional areas and other large urban centres was associated with longer 50^th^ and 75^th^ percentiles of time to diagnosis for hepatic and central nervous system (CNS) tumours. Diagnostic intervals were shorter in the post-pandemic period (2020–2022) across several cancer sites, with CNS tumours showing reductions across all quantiles.

**Interpretation:** Diagnostic timeliness differed by cancer type, age, and rurality, but not by sex, material, or social deprivation. The shorter diagnostic intervals observed in the post-pandemic period suggest that pandemic-related changes in care pathways may have expedited diagnosis for some cancers.

## Introduction

Timely diagnosis is a key tenet of high-quality cancer care as it improves the prospect of curable disease,^1^ reduces treatment burden, and lessens psychosocial distress for patients and their families.^2,3^ In adult populations, the importance of timely cancer care is well established and embedded within clinical pathways and screening programs for early detection.^4^ In contrast, for pediatric populations—where opportunity for early detection is often limited and presenting symptoms are often vague ^5-7^—the determinants and consequences of prolonged time to diagnosis are less well characterized. Understanding the factors that influence diagnostic timeliness is essential to ensuring equitable access and quality of care for children with cancer.

In Canada, research on diagnostic timeliness in childhood cancer has been predominantly descriptive,^8-10^ with limited analytic assessment of factors associated with its variation,^11^ and often restricted to single disease sites.^12-15^ Moreover, much of this work predates current diagnostic and treatment practices. The most recent pan-cancer estimates are more than 15 years old,^8^ and therefore do not capture recent system-level changes, including the impact of the COVID-19 pandemic.^16^ Contemporary evidence is needed to better understand the factors affecting time to diagnosis and ensure high-quality care for children and adolescents with cancer.

Our objective was to quantify the diagnostic interval in all children and adolescents with cancer in Québec, Canada diagnosed between 2010 and 2022, and to identify patient-, demographic-, and system-level predictors of time to diagnosis.

## Methods

### Study design and data sources

We conducted a retrospective, population-based cohort study using administrative health data from January 2010 to March 2022 in Québec, Canada. The province operates a publicly funded healthcare system that covers medically necessary services, defined as physician services and hospital care. We identified children and adolescents with at least one diagnostic code for malignant tumours between January 2012 and March 2022 in the *Maintenance et exploitation des données pour l’étude de la clientèle hospitalière* (MED-ECHO) dataset, which captures all hospitalizations and day surgeries.^17^ We classified cases into 12 diagnostic groups defined by the International Classification of Childhood Cancer, third edition (ICCC-3), using the published conversion algorithm to map International Classification of Diseases for Oncology, third edition (ICD-O-3) codes to ICCC-3 categories.^18^ Medical fee-for-service physician claims, diagnostic codes, and demographic information from the *Régie de l’assurance maladie du Québec* (RAMQ) were linked to MED-ECHO using unique identifiers. Data sources are described in Supplementary Table S1.

### Study population

Analyses were restricted to individuals insured under the public health plan, which covers 95% of the Québec population.^19,20^ We included children and adolescents aged 0–19 years at diagnosis with cancer. A two-year washout period (2010-2012) was applied to exclude prevalent cancer cases and ensure sufficient observation of healthcare utilization prior to diagnosis. Individuals aged 0–2 years diagnosed during the washout period were retained. To ensure case validity, we included only individuals with evidence of both a diagnostic (e.g., biopsy, imaging) and treatment encounter (e.g., surgery, chemotherapy, radiotherapy). We assigned the date of diagnosis hierarchically, prioritizing biopsy as the most definitive diagnostic procedure (Supplementary Table S2).

### Time to diagnosis

Our primary outcome was time to diagnosis, measured in days, from the first cancer-related healthcare encounter (i.e., index date) until diagnosis.^21^ Because time to diagnosis is not routinely captured in administrative health data, we determined the index healthcare encounter date using statistical process control charts.^22-24^ Control charts track encounter frequency to distinguish normal variation in healthcare use from increases that signal diagnostic activity. We identified the specialists, procedures, and diagnostic codes occurring more frequently in the 0–3 versus 18–24 months before diagnosis. These codes were grouped by clinical and functional similarity into encounter categories specific to each cancer type, broadly representing symptom-related encounters (linked to a diagnosis code) and procedure-based encounters (linked to an intervention code). We classified encounters as symptom-related if they included a control chart-identified diagnosis code on the index date. Billing codes for age-related encounters (e.g., immunizations) unrelated to cancer diagnosis were excluded. For each cancer-specific encounter category, statistical process control charts were used to determine the lookback period.^23^ Four cut-point rules were applied and signal strength was assessed.^24^ Categories with signal strength below 80% were excluded. For each patient, the earliest encounter among eligible cancer-related categories was assigned as the index date. When the index encounter was a procedure, we backdated the index date to the most recent visit with the referring provider in the last 12 months. Statistical process control charts, encounter categories, and lookback periods are further described in Supplementary Tables S3–S27.

Secondary outcomes included time to treatment (days), defined as the interval from diagnosis until initiation of treatment, and the total interval (days), defined as the time from the index date until initiation of treatment.^21^

### Exposures and covariates

We included demographic characteristics known to influence healthcare access, including age at index date (<1, 1–4, 5–9, 10–14, 15–19 years), sex, rurality (based on Statistics Canada Census Metropolitan Area classification), diagnosis period (pre-COVID-19: 2010– 2019; post-COVID-19: 2020–2022), and quintiles of the Material and Social Deprivation Index, assigned at the census dissemination area level using postal code at diagnosis.^25,26^

Additional binary variables were summarized descriptively at the index date, including presence of a predisposition syndrome, emergency department (ED) visit, any symptom, and imaging (Supplementary Table S28). Because these variables were derived from billing codes used to inform the outcome, we did not adjust for them in multivariable models.

### Statistical analysis

Diagnostic intervals were summarized as medians with interquartile ranges (IQR) for the overall cohort and by cancer type. Entry into the diagnostic pathway was summarized using mutually exclusive combinations of encounters present on the index date (ED visit, symptom-related, imaging) and stratified by disease group (leukemia/lymphomas, CNS tumours, solid tumours) and diagnosis period. Because time-to-diagnosis data are typically right-skewed, we used quantile regression to estimate differences in time to diagnosis at the 25^th^, 50^th^ (median), and 75^th^ percentiles.^27,28^ Multivariable models were adjusted for sex, age at index date, diagnosis period, and rurality, informed by evidence from existing literature.^8-10,29,30^ When covariate data were missing, we included a missing-indicator category to retain all individuals in the analysis. We evaluated potential effect modification using global Wald tests for interaction between cancer type and rurality, and between cancer type and diagnosis period.

### Ethics approval

This secondary data analysis was approved by the McGill Institutional Review Board, in accordance with the Canadian Tri-Council Policy Statement of Ethical Conduct for Research Involving Humans.

## Results

We identified 2927 children and adolescents diagnosed with cancer between 2010-2022 (Table 1). Demographic characteristics varied by cancer type, with younger ages predominating in leukemia, neuroblastoma, retinoblastoma, renal tumours, and hepatic tumours, and older ages in lymphoma, bone tumours, germ cell tumours, and carcinomas. Most patients resided in Montréal, and the distribution of material deprivation was similar across cancer types; overall, the largest proportion of cases was in the least socially deprived quintile. Missingness was low for rurality (0.92%) and both material and social deprivation (1.57%). Presentation through the ED at index and the presence of a predisposition syndrome were uncommon, whereas symptom-related encounters at index were frequent across cancer types.

**Table 1.**
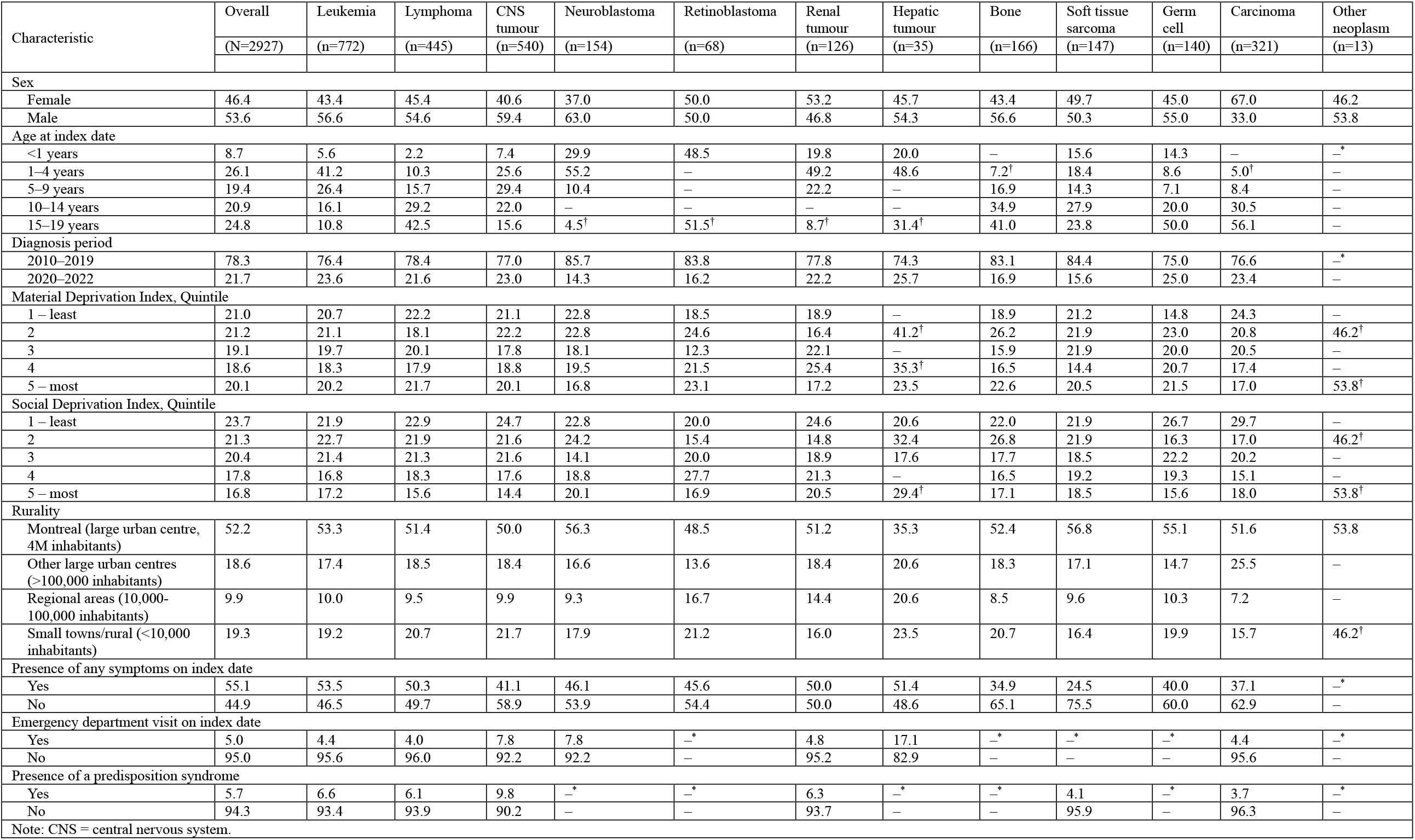
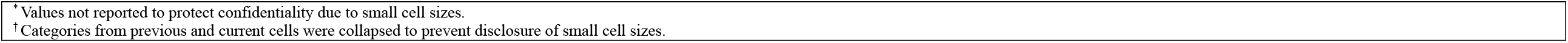
Demographic characteristics (%) of the study cohort, overall and by cancer type.

| Characteristic | Overall<br>(N=2927) | Leukemia<br>(n=772) | Lymphoma<br>(n=445) | CNS<br>tumour<br>(n=540) | Neuroblastoma<br>(n=154) | Retinoblastoma<br>(n=68) | Renal<br>tumour<br>(n=126) | Hepatic<br>tumour<br>(n=35) | Bone<br>(n=166) | Soft tissue<br>sarcoma<br>(n=147) | Germ<br>cell<br>(n=140) | Carcinoma<br>(n=321) | Other<br>neoplasm<br>(n=13) |
| --- | --- | --- | --- | --- | --- | --- | --- | --- | --- | --- | --- | --- | --- |
| Sex |  |  |  |  |  |  |  |  |  |  |  |  |  |
| Female | 46.4 | 43.4 | 45.4 | 40.6 | 37.0 | 50.0 | 53.2 | 45.7 | 43.4 | 49.7 | 45.0 | 67.0 | 46.2 |
| Male | 53.6 | 56.6 | 54.6 | 59.4 | 63.0 | 50.0 | 46.8 | 54.3 | 56.6 | 50.3 | 55.0 | 33.0 | 53.8 |
| Age at index date |  |  |  |  |  |  |  |  |  |  |  |  |  |
| <1 years | 8.7 | 5.6 | 2.2 | 7.4 | 29.9 | 48.5 | 19.8 | 20.0 | – | 15.6 | 14.3 | – | –* |
| 1–4 years | 26.1 | 41.2 | 10.3 | 25.6 | 55.2 | – | 49.2 | 48.6 | 7.2<sup>†</sup> | 18.4 | 8.6 | 5.0<sup>†</sup> | – |
| 5–9 years | 19.4 | 26.4 | 15.7 | 29.4 | 10.4 | – | 22.2 | – | 16.9 | 14.3 | 7.1 | 8.4 | – |
| 10–14 years | 20.9 | 16.1 | 29.2 | 22.0 | – | – | – | – | 34.9 | 27.9 | 20.0 | 30.5 | – |
| 15–19 years | 24.8 | 10.8 | 42.5 | 15.6 | 4.5<sup>†</sup> | 51.5<sup>†</sup> | 8.7<sup>†</sup> | 31.4<sup>†</sup> | 41.0 | 23.8 | 50.0 | 56.1 | – |
| Diagnosis period |  |  |  |  |  |  |  |  |  |  |  |  |  |
| 2010–2019 | 78.3 | 76.4 | 78.4 | 77.0 | 85.7 | 83.8 | 77.8 | 74.3 | 83.1 | 84.4 | 75.0 | 76.6 | –* |
| 2020–2022 | 21.7 | 23.6 | 21.6 | 23.0 | 14.3 | 16.2 | 22.2 | 25.7 | 16.9 | 15.6 | 25.0 | 23.4 | – |
| Material Deprivation Index, Quintile |  |  |  |  |  |  |  |  |  |  |  |  |  |
| 1 – least | 21.0 | 20.7 | 22.2 | 21.1 | 22.8 | 18.5 | 18.9 | – | 18.9 | 21.2 | 14.8 | 24.3 | – |
| 2 | 21.2 | 21.1 | 18.1 | 22.2 | 22.8 | 24.6 | 16.4 | 41.2<sup>†</sup> | 26.2 | 21.9 | 23.0 | 20.8 | 46.2<sup>†</sup> |
| 3 | 19.1 | 19.7 | 20.1 | 17.8 | 18.1 | 12.3 | 22.1 | – | 15.9 | 21.9 | 20.0 | 20.5 | – |
| 4 | 18.6 | 18.3 | 17.9 | 18.8 | 19.5 | 21.5 | 25.4 | 35.3<sup>†</sup> | 16.5 | 14.4 | 20.7 | 17.4 | – |
| 5 – most | 20.1 | 20.2 | 21.7 | 20.1 | 16.8 | 23.1 | 17.2 | 23.5 | 22.6 | 20.5 | 21.5 | 17.0 | 53.8<sup>†</sup> |
| Social Deprivation Index, Quintile |  |  |  |  |  |  |  |  |  |  |  |  |  |
| 1 – least | 23.7 | 21.9 | 22.9 | 24.7 | 22.8 | 20.0 | 24.6 | 20.6 | 22.0 | 21.9 | 26.7 | 29.7 | – |
| 2 | 21.3 | 22.7 | 21.9 | 21.6 | 24.2 | 15.4 | 14.8 | 32.4 | 26.8 | 21.9 | 16.3 | 17.0 | 46.2<sup>†</sup> |
| 3 | 20.4 | 21.4 | 21.3 | 21.6 | 14.1 | 20.0 | 18.9 | 17.6 | 17.7 | 18.5 | 22.2 | 20.2 | – |
| 4 | 17.8 | 16.8 | 18.3 | 17.6 | 18.8 | 27.7 | 21.3 | – | 16.5 | 19.2 | 19.3 | 15.1 | – |
| 5 – most | 16.8 | 17.2 | 15.6 | 14.4 | 20.1 | 16.9 | 20.5 | 29.4<sup>†</sup> | 17.1 | 18.5 | 15.6 | 18.0 | 53.8<sup>†</sup> |
| Rurality |  |  |  |  |  |  |  |  |  |  |  |  |  |
| Montreal (large urban centre, 4M inhabitants) | 52.2 | 53.3 | 51.4 | 50.0 | 56.3 | 48.5 | 51.2 | 35.3 | 52.4 | 56.8 | 55.1 | 51.6 | 53.8 |
| Other large urban centres (>100,000 inhabitants) | 18.6 | 17.4 | 18.5 | 18.4 | 16.6 | 13.6 | 18.4 | 20.6 | 18.3 | 17.1 | 14.7 | 25.5 | – |
| Regional areas (10,000–100,000 inhabitants) | 9.9 | 10.0 | 9.5 | 9.9 | 9.3 | 16.7 | 14.4 | 20.6 | 8.5 | 9.6 | 10.3 | 7.2 | – |
| Small towns/rural (<10,000 inhabitants) | 19.3 | 19.2 | 20.7 | 21.7 | 17.9 | 21.2 | 16.0 | 23.5 | 20.7 | 16.4 | 19.9 | 15.7 | 46.2<sup>†</sup> |
| Presence of any symptoms on index date |  |  |  |  |  |  |  |  |  |  |  |  |  |
| Yes | 55.1 | 53.5 | 50.3 | 41.1 | 46.1 | 45.6 | 50.0 | 51.4 | 34.9 | 24.5 | 40.0 | 37.1 | –* |
| No | 44.9 | 46.5 | 49.7 | 58.9 | 53.9 | 54.4 | 50.0 | 48.6 | 65.1 | 75.5 | 60.0 | 62.9 | – |
| Emergency department visit on index date |  |  |  |  |  |  |  |  |  |  |  |  |  |
| Yes | 5.0 | 4.4 | 4.0 | 7.8 | 7.8 | –* | 4.8 | 17.1 | –* | –* | –* | 4.4 | –* |
| No | 95.0 | 95.6 | 96.0 | 92.2 | 92.2 | – | 95.2 | 82.9 | – | – | – | 95.6 | – |
| Presence of a predisposition syndrome |  |  |  |  |  |  |  |  |  |  |  |  |  |
| Yes | 5.7 | 6.6 | 6.1 | 9.8 | –* | –* | 6.3 | –* | –* | 4.1 | –* | 3.7 | –* |
| No | 94.3 | 93.4 | 93.9 | 90.2 | – | – | 93.7 | – | – | 95.9 | – | 96.3 | – |

Note: CNS = central nervous system.
\*Values not reported to protect confidentiality due to small cell sizes.
†Categories from previous and current cells were collapsed to prevent disclosure of small cell sizes.

Median time to diagnosis varied by cancer type, ranging from 8 days for leukemia, 8 days for renal tumours, and 8 days for hepatic tumours, to 41 days for soft tissue sarcomas and 100 days for carcinomas. Older children had longer time to diagnosis, with those aged 15–19 years having a median of 36 days. Medians were similar across levels of material and social deprivation and by rurality (Table 2). Individuals diagnosed during the post-COVID-19 period (2020–2022) had shorter median diagnostic intervals (14 days) than those diagnosed during 2010–2019 (22 days). Median time to treatment and the total interval varied minimally by covariates, with the longest total intervals observed for adolescents (15–19 years, 42 days) and patients with carcinomas (100 days) (Supplementary Table S29). Entry into the diagnostic pathway varied by cancer type. CNS tumours accounted for the highest proportion of ED presentations, whereas most non-ED presentations were non-symptom-related. Leukemias and lymphomas were predominantly symptom-related without imaging, whereas solid tumours were primarily non-symptom-related without imaging. In the post-pandemic period, ED presentations increased, particularly among CNS tumours, and symptom-related cases without imaging declined across cancer types (Figure 1A–C). In univariate quantile regression, differences in time to diagnosis were most pronounced by age and cancer type at the 75^th^ percentile. Time to diagnosis showed a broad distribution for cancers overall and a bimodal distribution for CNS tumours and leukemia and lymphoma combined (Figure 2).

**Table 2.** Median and interquartile range of the diagnostic interval and univariate quantile regression of time to diagnosis.

| Characteristic | Observed Diagnosis interval (days) |  | Modeled difference in diagnostic interval (days) |  |  |  |  |  |
| --- | --- | --- | --- | --- | --- | --- | --- | --- |
|  | Median | IQR | 25<sup>th</sup> percentile | CI (95%) | 50<sup>th</sup> percentile | CI (95%) | 75<sup>th</sup> percentile | CI (95%) |
| Sex |  |  |  |  |  |  |  |  |
| Female | 23 | (5 to 82) | 0 | (-1 to 1) | 5 | (-0 to 10) | 11 | (-3 to 25) |
| Male | 18 | (5 to 72) | Ref. | Ref. | Ref. | Ref. | Ref. | Ref. |
| Age group at index |  |  |  |  |  |  |  |  |
| <1 year | 18 | (5 to 69) | 1 | (-0 to 2) | 10 | (2 to 18) | 38 | (4 to 72) |
| 1-4 years | 13 | (4 to 69) | Ref. | Ref. | Ref. | Ref. | Ref. | Ref. |
| 5-9 years | 15 | (4 to 61) | 1 | (-1 to 1) | 1 | (-4 to 6) | 1 | (-17 to 19) |
| 10-14 years | 20 | (5 to 70) | 1 | (-0 to 2) | 8 | (2 to 14) | 15 | (-7 to 37) |
| 15-19 years | 36 | (8 to 109) | 4 | (3 to 5) | 19 | (12 to 26) | 39 | (19 to 59) |
| Cancer type |  |  |  |  |  |  |  |  |
| Leukemia | 8 | (3 to 44) | Ref. | Ref. | Ref. | Ref. | Ref. | Ref. |
| Lymphoma | 25 | (8 to 60) | 5 | (4 to 6) | 18 | (13 to 23) | 20 | (9 to 31) |
| CNS tumour | 28 | (7 to 129) | 4 | (3 to 5) | 21 | (14 to 28) | 96 | (65 to 127) |
| Neuroblastoma | 11 | (4 to 45) | 1 | (-0 to 2) | 3 | (-3 to 9) | 2 | (-16 to 20) |
| Retinoblastoma | 9 | (3 to 51) | 0 | (-1 to 1) | 1 | (-6 to 8) | 6 | (-46 to 58) |
| Renal tumour | 8 | (5 to 15) | 2 | (1 to 3) | 0 | (-4 to 4) | -13 | (-35 to 9) |
| Hepatic tumour | 8 | (4 to 19) | 1 | (-0 to 2) | 0 | (-7 to 7) | -25 | (-60 to 9) |
| Bone | 15 | (8 to 43) | 5 | (3 to 7) | 7 | (2 to 12) | 3 | (-11 to 17) |
| Sarcoma | 41 | (11 to 151) | 8 | (5 to 11) | 36 | (22 to 50) | 125 | (69 to 181) |
| Germ cell | 15 | (6 to 28) | 2 | (0 to 4) | 7 | (2 to 12) | -11 | (-23 to 1) |
| Carcinoma | 100 | (33 to 192) | 30 | (24 to 36) | 101 | (85 to 117) | 163 | (140 to 186) |
| Other neoplasm | 9 | (2 to 17) | 1 | (-9 to 11) | 2 | (-7 to 11) | -25 | (-193 to 143) |
| Diagnosis period |  |  |  |  |  |  |  |  |
| 2010-2019 | 22 | (5 to 79) | Ref. | Ref. | Ref. | Ref. | Ref. | Ref. |
| 2020-2022 | 14 | (4 to 70) | -1 | (-2 to 0) | -8 | (-12 to -4) | -10 | (-26 to 6) |
| Material Deprivation Index, Quintile |  |  |  |  |  |  |  |  |
| 1 – least | 23 | (5 to 79) | Ref. | Ref. | Ref. | Ref. | Ref. | Ref. |
| 2 | 19 | (5 to 78) | 0 | (-2 to 2) | -4 | (-11 to 3) | -1 | (-23 to 21) |
| 3 | 20 | (5 to 81) | 0 | (-2 to 2) | -3 | (-9 to 3) | 2 | (-23 to 27) |
| 4 | 22 | (5 to 78) | 0 | (-2 to 2) | -1 | (-8 to 6) | -1 | (-23 to 21) |
| 5 – most | 18 | (4 to 74) | 0 | (-2 to 0) | -6 | (-13 to 1) | -5 | (-25 to 15) |
| Social Deprivation Index, Quintile |  |  |  |  |  |  |  |  |
| 1 – least | 23 | (5 to 81) | Ref. | Ref. | Ref. | Ref. | Ref. | Ref. |
| 2 | 21 | (5 to 78) | 0 | (-1 to 1) | -2 | (-8 to 4) | -2 | (-23 to 19) |
| 3 | 17 | (5 to 69) | 0 | (-2 to 2) | -6 | (-12 to 0) | -11 | (-28 to 6) |
| 4 | 21 | (5 to 82) | 0 | (-2 to 2) | -2 | (-9 to 5) | 2 | (-19 to 23) |
| 5 – most | 17 | (4 to 78) | -1 | (-2 to 0) | -6 | (-13 to 1) | -2 | (-26 to 22) |
| Rurality |  |  |  |  |  |  |  |  |
| Montreal (large urban centre, 4M inhabitants) | 20 | (5 to 75) | Ref. | Ref. | Ref. | Ref. | Ref. | Ref. |
| Other large urban centres (>100,000 inhabitants) | 21 | (5 to 78) | 0 | (-1 to 1) | 1 | (-4 to 6) | 3 | (-17 to 23) |
| Regional areas (10,000-100,000 inhabitants) | 22 | (6 to 92) | 1 | (-1 to 3) | 1 | (-9 to 11) | 17 | (-15 to 49) |
| Small towns/rural (<10,000 inhabitants) | 19 | (5 to 79) | 0 | (-1 to 1) | -2 | (-7 to 3) | 4 | (-11 to 19) |

Note: CI = confidence interval; CNS = central nervous system; IQR = interquartile range; Ref. = reference category

**Figure 1.**
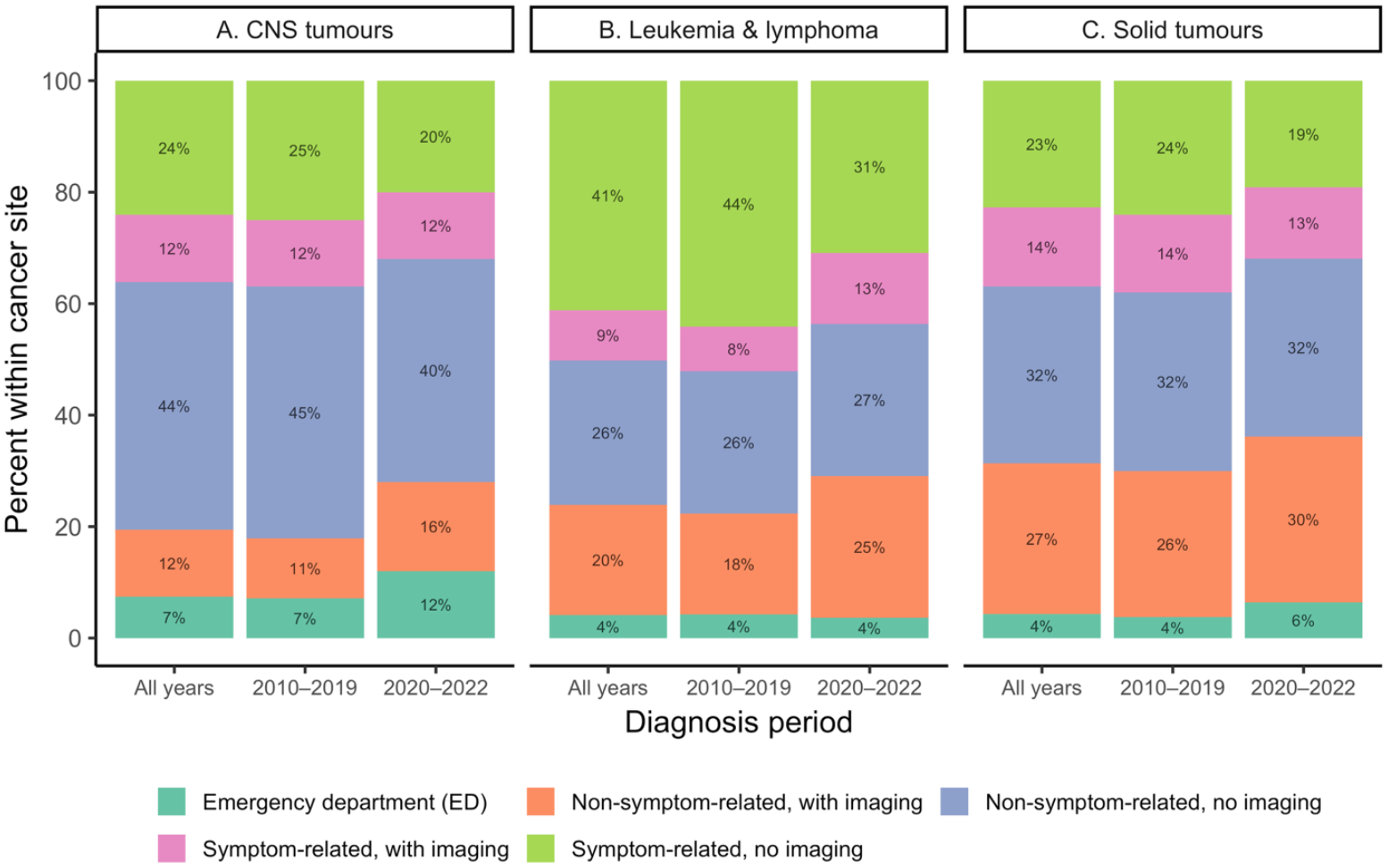
Entry into the diagnostic pathway by cancer site and diagnosis period. Percentage of encounters recorded on the index date by time period, shown separately for central nervous system (CNS) tumours, leukemias/lymphomas, and solid tumours. Stacked bars indicate the proportion occurring in all years, pre-pandemic (2010–2019) and post-pandemic (2020–2022).

**Figure 2.**
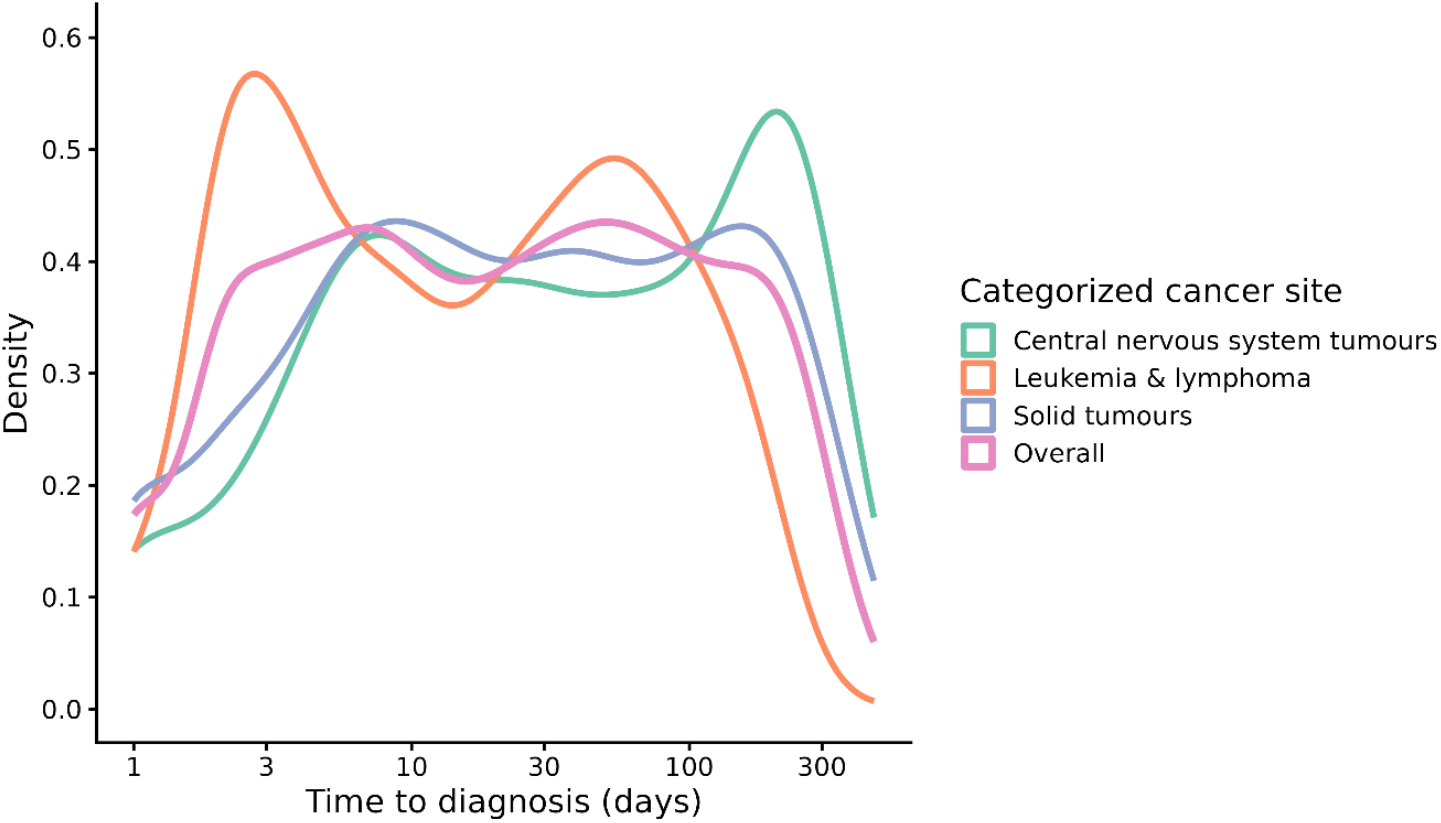
Distribution of time to diagnosis (log scale), overall and by broad cancer site. Kernel density plot of log-transformed time to diagnosis (days). Distributions are shown for cancers overall and by categorized cancer site (central nervous system tumours, leukemias and lymphomas, solid tumours). Log transformation was applied to accommodate skewness and allow comparison of distributions across cancer sites.

Table 3 presents adjusted quantile regression models evaluating the 25^th^, 50^th^, and 75^th^ percentiles of time to diagnosis stratified by cancer type. Models were not fitted for the cancer type “other neoplasms” due to small numbers. Sex was not associated with time to diagnosis across cancer types. Age patterns differed by cancer type and percentile: among CNS tumours, infants (<1 year) at the 75^th^ percentile of time to diagnosis waited 81 days longer than 1–4-year-olds at the same percentile, whereas for sarcomas, 5–9-year-olds had 167-day shorter intervals at the 75^th^ percentile compared to 10–14-year-olds. Shorter time to diagnosis was also observed among younger children with leukemia, renal tumours, germ cell tumours, and carcinomas, although the strength and presence of the association differed across percentiles. During 2020–2022, diagnostic intervals were notably shorter for CNS tumours across all quantiles compared to 2010–2019, with a 46-day reduction at the 75^th^ percentile compared with 2010-2019. By rurality, children with CNS tumours living in regional areas had diagnostic intervals 91 days longer at the 75^th^ percentile than those residing in Montréal. At the median, children with hepatic tumours living in regional areas waited 30 days longer for diagnosis than their urban counterparts. For bone tumours, children residing in other large urban centres waited 18 days longer for diagnosis compared to those living in Montréal.

**Table 3.**
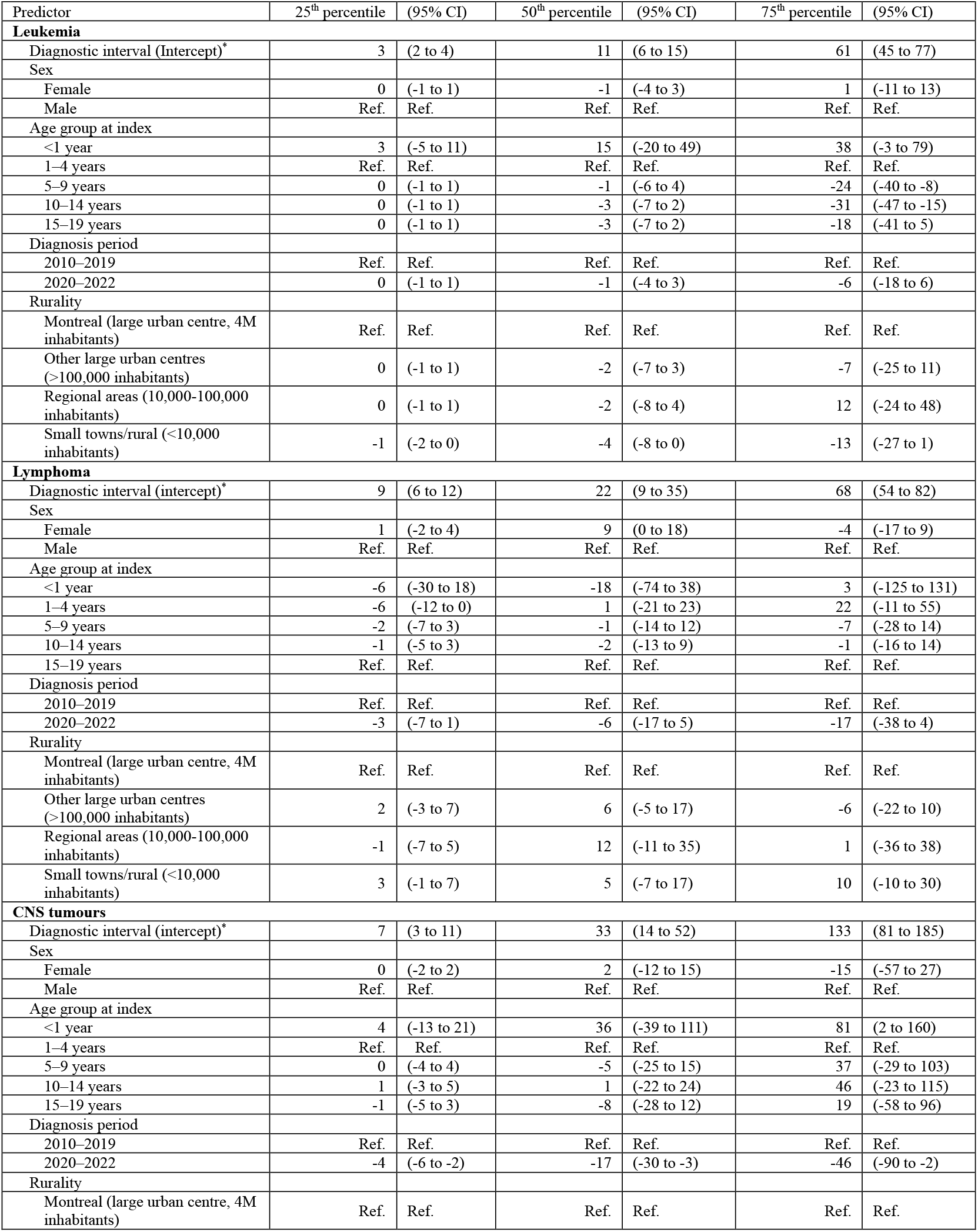
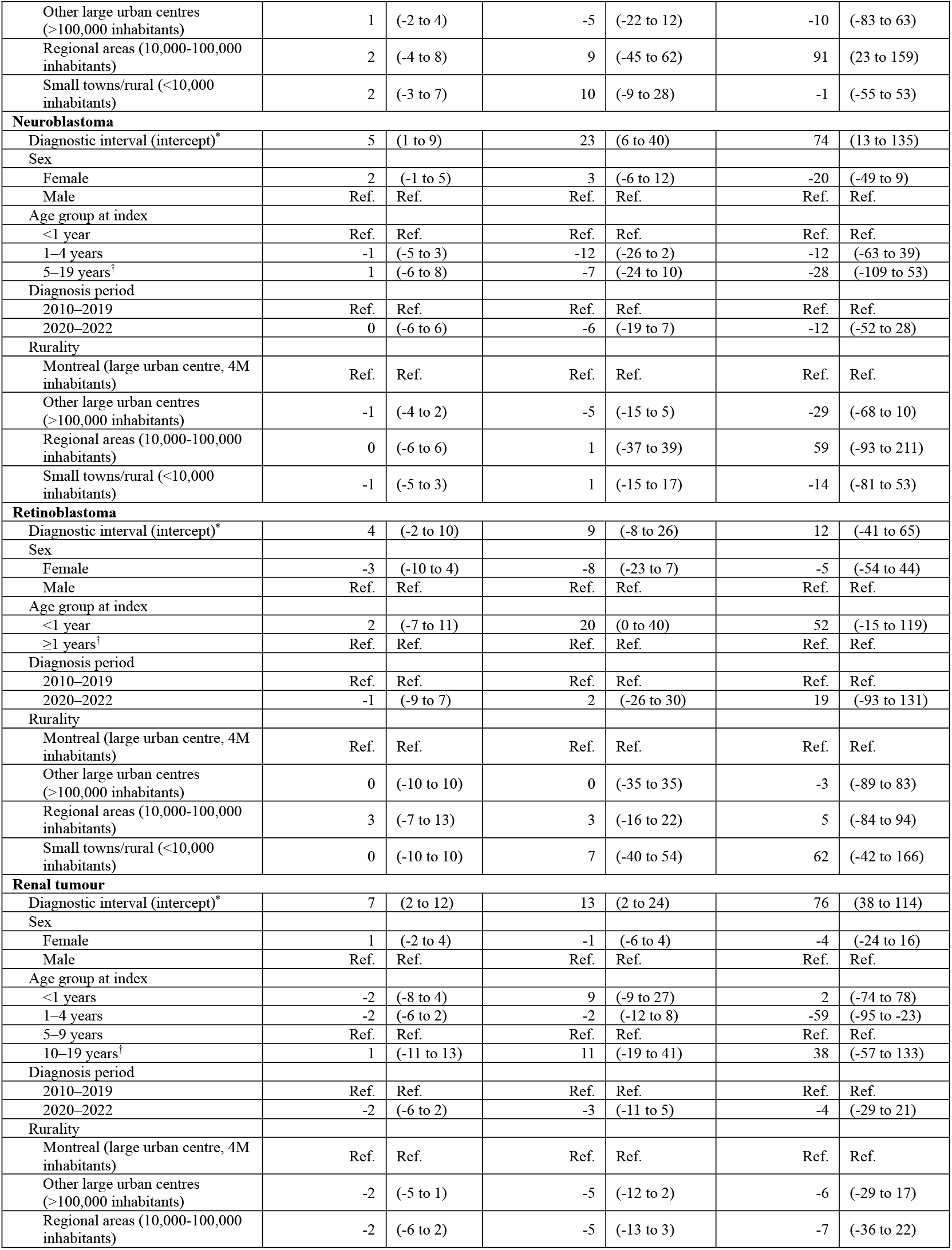
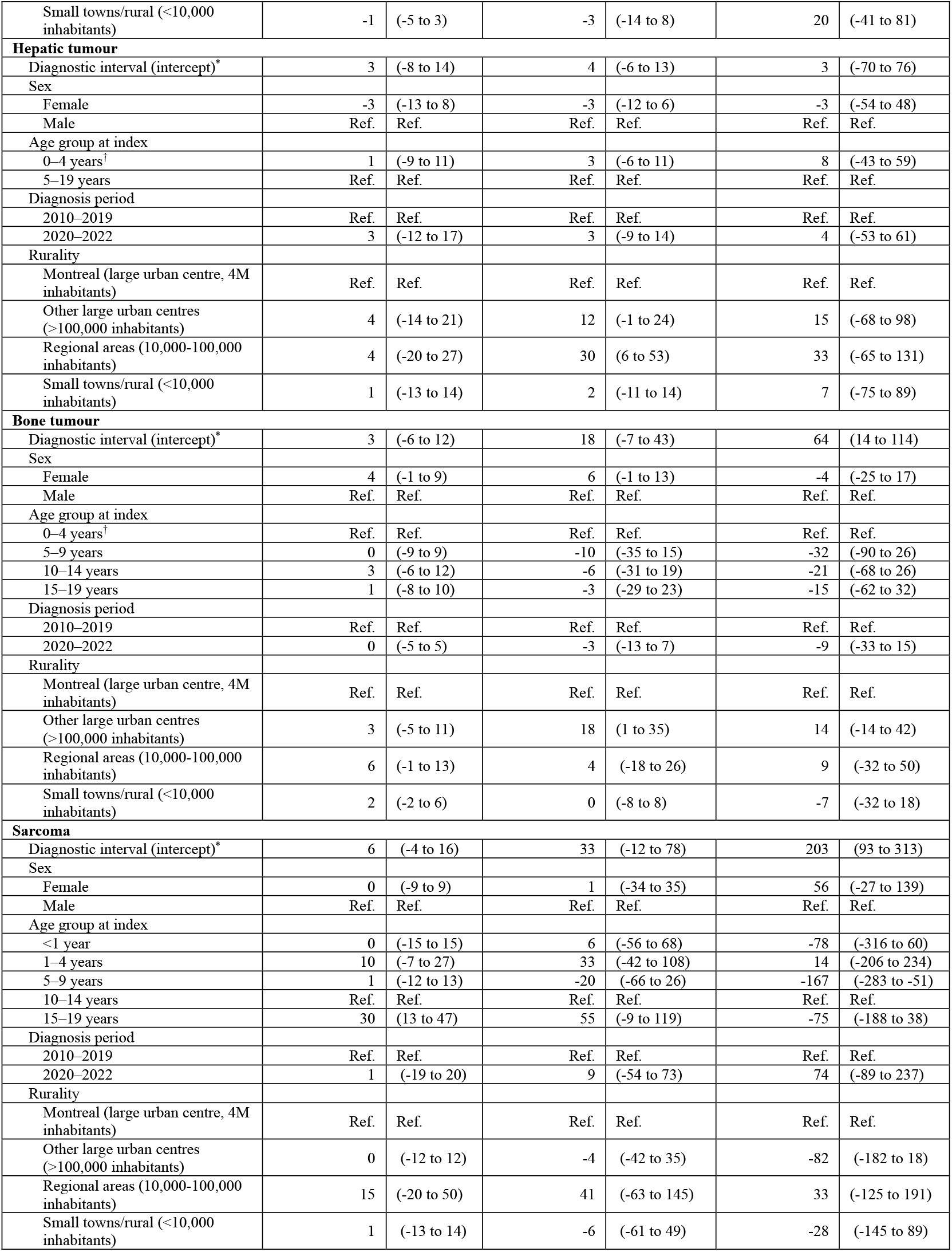
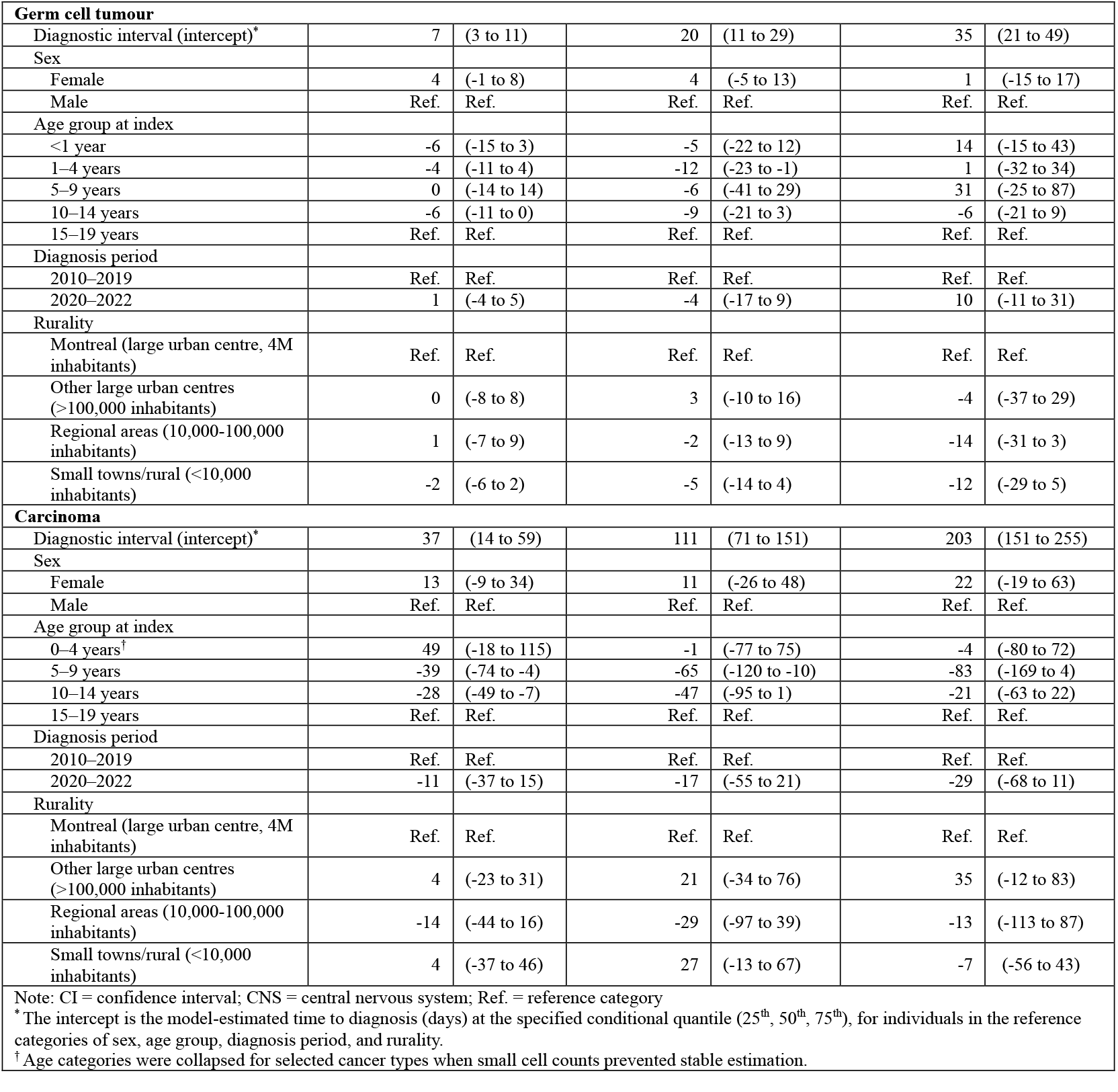
Adjusted diagnostic interval estimates (days) from multivariable quantile regression models, stratified by cancer type.

There was no evidence of interaction between cancer type and rurality across quantiles. Evidence of interaction between cancer type and diagnosis period was observed at the 25^th^ percentile (p=0.038), but not at the 50^th^ or 75^th^ percentiles.

### Interpretation

We found that diagnostic intervals varied substantially by cancer type for childhood and adolescent cancers diagnosed in Québec between 2010-2022. Overall, we did not observe significant differences by sex, material and social deprivation quintiles, or by rurality overall. However, we did observe longer times to diagnosis for specific cancer types by rurality and with increasing age, and shorter times in the post-pandemic period (2020-2022). Quantile regression revealed that differences were magnified at higher percentiles, suggesting prolonged time to diagnosis is concentrated within a smaller subset of patients.

Our findings are consistent with those reported in Canada and other regions, which have identified longer diagnostic intervals in older children.^8,9,30-33^ This pattern may reflect developmental factors such as increasing patient autonomy and challenges recognizing signs and symptoms,^34,35^ as well as differences in disease types and diagnostic pathways,^36^ given that cancers more common in adolescence—including bone tumours, lymphomas, and germ cell tumors^37^—often require multiple investigations before diagnosis. Differences by cancer type also followed expected clinical patterns: cancers with more acute and specific presentations, such as leukemias,^34,38^ had shorter diagnostic intervals, whereas carcinomas, which encompass thyroid cancers and melanomas, showed the longest intervals, likely reflecting the monitoring of indolent or nonspecific findings prior to disease confirmation.^33,39,40^

We observed limited association between diagnostic timeliness and area-level material and social deprivation, and moderate variation by rurality. The largest geographic difference occurred among CNS tumours, where, at the 75^th^ percentile, children residing in regional areas waited three months longer than those in urban centres. Time to diagnosis at the 50^th^ percentile was longer for children with bone tumours residing in other large urban centres and for those with hepatic tumours residing in regional areas. These differences may reflect diagnostic challenges related to referral patterns and locus of care, as children with these diseases are often evaluated in adult facilities where suspicion of cancer may be lower.^36,41,42^ Heterogeneity in tumour subtype and the concentration of diagnostic services in urban tertiary care facilities may further contribute to these differences.^29,43,44^ Although Canadian studies examining median diagnostic intervals have reported no overall urban-rural or sociodemographic differences,^9,13,14,45^ our findings indicate that disparities may exist for specific cancers or segments of the distribution of time to diagnosis.

Interestingly, diagnostic intervals were shorter in 2020–2022 compared with 2010–2019, suggesting pandemic-related shifts in healthcare-seeking behaviour and care delivery. In Canada, childhood cancer incidence remained stable during the early pandemic period,^46^ reducing the likelihood that our findings reflect incomplete case ascertainment. Interaction between cancer type and diagnosis period at the 25^th^ percentile indicates that changes primarily affected children diagnosed most quickly. Consistent with this, ED-initiated diagnostic pathways increased during 2020–2022, particularly for CNS tumours, suggesting that higher-acuity presentations or altered health-seeking behaviour contributed to expedited diagnostic evaluation.^47,48^ Although pediatric ED volumes decreased in Canada during the pandemic, visits that occurred were of higher acuity than in the pre-pandemic period.^49,50^ Evidence from adult oncology indicates increased peri-diagnostic ED use during the pandemic, concordant with a shift toward ED-initiated diagnostic pathways.^51^ In non-ED pathways, presentations without imaging declined while imaging-based presentations increased, suggesting consolidation of diagnostic investigations under COVID-19 clinical practice guidelines.^52^

Evidence from adult populations has linked prolonged time to diagnosis with adverse outcomes across multiple cancers,^2,53-56^ suggesting that clinically meaningful effects may be concentrated among individuals with the longest diagnostic intervals. Although median time to diagnosis was short in our pediatric cohort, a subset experienced prolonged intervals, underscoring the importance of examining the distribution of time to diagnosis even when average values seem favourable. Quantile regression estimates associations at user-specified percentiles, enabling direct evaluation of the determinants of prolonged diagnostic intervals and identification of subgroups most likely to benefit from earlier cancer recognition. Future work should extend this framework to larger, multi-jurisdictional datasets with richer clinical data to examine additional clinical and contextual determinants of longer time to diagnosis.

### Limitations

We used administrative health data to derive outcome measures as direct assessment was not possible. These data may miss relevant diagnostic encounters, particularly for patients with multiple or nonspecific complaints,^19^ and without patient-reported symptom onset, the diagnostic interval is likely underestimated. For individuals whose index encounter was a procedure, the referral date was used when available; missing referral dates (5.91% overall) may contribute to underestimated diagnostic intervals. Cancer type was classified using a validated algorithm that accurately identifies major ICCC-3 groups but has documented subtype misclassification,^57^ limiting finer stratification and potentially obscuring within-group heterogeneity. Finally, the rarity of childhood cancer produced small numbers in some disease and covariate strata, reducing precision and limiting power to detect clinically meaningful differences in time to diagnosis across cancer types. Although we observed interaction between cancer type and diagnosis period, the specific cancers driving this effect could not be determined with confidence.

### Conclusion

Diagnostic intervals were longer for older children and for those with sarcomas and carcinomas, and shorter during 2020–2022. Quantile regression revealed that the longest diagnostic intervals were concentrated among children with CNS, hepatic, and bone tumours, with impacts varying by age and rurality. Efforts to improve diagnostic timeliness must address both tumour-specific diagnostic challenges and health-system barriers, including point of entry to care and proximity to tertiary facilities, to reduce inequities in time to diagnosis.

## Data Availability

This study used data from the © Government of Quebec (2022) under licence from the Institut de la statistique du Quebec. The Government of Quebec is not responsible for the analyses, compilations, interpretations, or conclusions presented in this manuscript. Restrictions apply to the availability of these data, which are regulated by Quebec law. Access can be requested by eligible researchers via application through Data Access Services (https://statistique.quebec.ca/en/services-recherche/submitting-a-
request). Codes for the analyses reported in this manuscript are publicly available on the Borealis repository at https://doi.org/10.5683/SP3/QISKU2.

https://statistique.quebec.ca/en/services-recherche/submitting-a-request

https://doi.org/10.5683/SP3/QISKU2

## Contributors

All authors were involved in the study conceptualization. Callum Mullen, Talía Malagón, Erin Strumpf, and Eduardo Franco developed the data analysis plan. Callum Mullen conducted the analyses and drafted the manuscript. All authors contributed to data interpretation, revised the manuscript for intellectual content, and gave final approval of the version to be published. All authors agreed to be accountable for all aspects of the work.

## Data sharing

This study used data from the © Government of Québec (2022) under licence from the Institut de la statistique du Québec. The Government of Québec is not responsible for the analyses, compilations, interpretations, or conclusions presented in this manuscript. Restrictions apply to the availability of these data, which are regulated by Québec law. Access can be requested by eligible researchers via application through Data Access Services (https://statistique.quebec.ca/en/services-recherche/submitting-a-request). Codes for the analyses reported in this manuscript are publicly available on the Borealis repository at https://doi.org/10.5683/SP3/QISKU2.

## Acknowledgements

We gratefully acknowledge Sarah Botting-Provost and Rodrigo Noorani from the Division of Cancer Epidemiology at McGill University for their contributions to the development of the codebooks (translation from French to English) and the maintenance of documentation for data used in this study. We also thank Laura E. Davis from McGill University for their substantive expertise and analytic guidance.

