## Supplementary Material for "Time to diagnosis among children and adolescents with cancer in Quebec, Canada: a population-based study"

**Table of Contents**

Supplementary Table S1. Description of data sources and study variables ……………….……… 2

Supplementary Table S2. Hierarchical assignment of the date of diagnosis ……………………. 3

Supplementary Table S3. Steps in applying statistical process control charts ……….………. 3–4

Supplementary Tables S4–S15. Cancer-specific encounter categories ……………….….… 5–21

Supplementary Tables S16–S27. Encounter category signal strength and lookback window derived from control charts for each cancer type ……………………………..….… 22–37

Supplementary Table S28. Derivation of binary variables used to describe index encounters and entry into the diagnostic pathway …………..…………………….……………………… 38

Supplementary Table S29. Median and interquartile range of the treatment interval and total interval …….……...………….…………………….……………………….…………….… 39

Supplementary References ……..………………….………………….……………….……… 40

Supplementary Table S1. Description of data sources and study variables

| **Database** | **Dataset** | **Variable** | **Description** |
|---|---|---|---|
| *Ministère de la Santé et des Services Sociaux;* MED-ECHO | Diagnosis file | Diagnosis | Medical diagnosis according to ICD-9/10. |
| | Service files | Specialization | Code indicating the specialization of the service provider. |
| | Intervention file | Date of intervention | Date the medical intervention was conducted. |
| | | Intervention code | Medical intervention code according to the Canadian Classification of Health Interventions (CCI). |
| | | Specialization of clinician performing intervention | Indicator for the specialization of the physician or provider who delivered the intervention. |
| | Tumour file | Cancer diagnosis | Diagnosis code indicating the tumor site, according to ICD-10. |
| | | Histology | Identification of the histological type of the tumour according to ICD-O-3. |
| *Régie de l'assurance maladie du Québec;* RAMQ | Registration file | Date of birth | Birth date of the insurant. |
| | | Sex | Sex of the insurant |
| | Services file | Prescriber ID | Unique ID of the healthcare professional who submitted the claim. |
| | | Prescriber specialization | Specialty code of the healthcare provider performing the service. |
| | | Service code | Fee-for-service remuneration code for procedures or actions performed by clinicians (*rémunération à l'acte*). |
| | | Date of service | Date the service was delivered. |
| | | Referring physician specialization | Medical specialization of the referring healthcare provider. |
| | | Referring physician ID | Unique ID of the referring healthcare provider. |
| | Diagnosis file | Diagnosis | Medical diagnosis according to ICD-10. |
| | Deprivation index file | Rurality | Rurality defined using Statistics Canada Census Metropolitan Area categories. |
| | | Material deprivation index | Quintile of material components. Area-based measure of material deprivation derived from census data, incorporating proportions of persons without a high school diploma, employment-to-population ratio, and average income within census areas.[1,2] |
| | | Social deprivation index | Quintile of social components. Area-based measure of social deprivation derived from census data, incorporating proportions of persons living alone, single-parent families, and individuals who are separated, divorced, or widowed.[1,2] |
| | | Reference date | Date of postal code record, indicating which census cycle was used to assign rurality and deprivation measures. |

NOTE: each data file contains the insurant specific unique ID allowing linkage between datasets.

| Supplementary Table S2. Hierarchical assignment of the date of diagnosis | | | |
|---|---|---|---|
| **Priority** | **Diagnostic Event** | **Operational Definition** | **Data Source** |
| 1 | Biopsy | Date of first histologic confirmation of malignancy through tissue biopsy (incisional, needle, core, excisional) recorded in MED-ECHO or RAMQ. | CCI codes: All diagnostic procedures with intervention type 71 (biopsy) or 87 (excision, including excisional biopsy), across all anatomical groups (e.g., 2.AA.71.^^, 2.YA.71.^^). Wildcard suffix ^^ denotes inclusion of all qualifier variants.<br><br>ACTE codes: Biopsy procedures identified in RAMQ physician billing data were included. Because ACTE is a non-hierarchical billing nomenclature with a large number of site- and modality-specific procedure codes, we do not reproduce the exhaustive code list. Biopsy was defined algorithmically by selection all ACTE codes whose official RAMQ descriptors contained biopsy or related terms. |
| 2 | Imaging | Earliest date of imaging procedure specific to cancer site, used only when no biopsy is available (CT, MRI, PET/CT, ultrasound and X-ray considered for select cancers). | CCI codes: All imaging procedures with intervention type 20 (CT scan), 40 (MRI), 70 (nuclear imaging, including PET), 10 (X-ray), and 30 (ultrasound) across all anatomical sites (e.g., 3.AN.20^^, 3.ZZ.70^^). Wildcard suffix ^^ denotes inclusion of all qualifier variants.<br><br>ACTE codes: Imaging procedures identified in RAMQ physician billing data were included. Because ACTE is a non-hierarchical billing nomenclature with a large number of site- and modality-specific codes, we do not reproduce the exhaustive code list. Imaging was defined algorithmically using all ACTE codes containing the keyword MRI, CT, PET, X-ray, ultrasound or related descriptors in the RAMQ billing nomenclature. |
| 3 | Oncology consultation / hospital visit | Earliest visit with an oncology specialist or cancer-related hospital admission. | ACTE codes: Oncology consultations were identified using RAMQ physician billing data corresponding to hematology/oncology consultation or assessments billed by a specialist in hematology/oncology. ACTE codes used include: 15005, 15007, 15020, 15021, 15122, 15058, 9150, 9160, 9162, 9170. |

| Supplemental Table S3. Detailed description of steps followed in determining the diagnostic interval using statistical process control charts | | | |
|---|---|---|---|
| **Steps** | | **Description** | **Notes** |
| 1 | Identify relevant physician specialties | Determine which physician specialties are seen more frequently by patients in the 0-3 months before diagnosis compared to the control period. | The control period was defined as the 18-24 months before diagnosis, when healthcare encounters are not likely to be related to underlying cancer. We identified the specialties relevant for each cancer type. |
| 2 | Identify and categorize cancer-related healthcare encounters | Among the physician specializations identified in step 1, identify the diagnoses and procedures performed more often in the 0-3 months before diagnosis compared to the control period. | Billing codes were identified in both RAMQ and MED-ECHO datasets. Codes are reported as CCI, ACTE, and ICD-9/10. We performed this step separately for each cancer type. |
| | | Using the encounter codes identified, create groups of cancer-related encounters for diagnostic and procedure codes, grouping codes based on similarity. | Cancer-related encounter categories were developed for each cancer type in consultation with a paediatric oncologist. |

| | | | |
|---|---|---|---|
| 3 | Plot statistical process control charts to derive cancer-specific lookback periods for each encounter category | In both the diagnosis period and control periods, calculate the weekly counts for each encounter category, and then compute the mean and standard deviation of these weekly counts. | The diagnosis period is the 0-18 months before diagnosis and the control period (or background period) is the 18-24 months before diagnosis. we calculate these values for each cancer. |
| | | Apply control chart rules 1-4 at each week leading to diagnosis. The lookback period stops when rules can no longer be applied. | As rules are applied, increasingly shorter lookback periods are identified. If few events occur in a week, rules will not be triggered. |
| | | | **Rule 4**: eight successive weekly counts above the mean background encounter rate.<br>**Rule 3:** four of five successive weekly counts more than one standard deviation above the mean background encounter rate.<br>**Rule 2**: two of three successive weekly counts more than two standard deviation above the mean background encounter rate.<br>**Rule 1:** a weekly count greater than or equal to three standard deviations above the mean background encounter rate. |
| 4 | Determine the signal strength for each encounter category | Discard any encounter categories that do not reach signal strength cutoff. | Signal strength represents the proportion of encounters within the lookback period that were above the expected number based on the background rate. Signal strength is calculated as: (A/C) *100, where C represents the total number of encounters within a lookback period; B is the estimated number of encounters in the lookback period that are not attributable to cancer diagnosis (i.e, background medical encounters). A is an estimate of the number of encounters in the lookback period that can be attributed to childhood cancer care, calculated as A = C – B. A signal strength cutoff of 80% was used.<br><br>Because few cases of the “other neoplasm” cancer type were observed, signal strength was unreliable. Rather than applying a signal strength cutoff, signals were treated as flags, and the first rule to flag was used as the lookback period. This applied to <1% of the total cohort. |
| 5 | Identify all the eligible encounters | For each individual, identify all cancer-specific healthcare encounters occurring within the defined lookback periods. | When RAMQ and MED-ECHO list encounters on the same day, give priority to the MED-ECHO record because it is validated by medical archivists.[3] |
| 6 | Add the referral date for procedure-based index events | Look back 12 months to identify the most proximal visit to the physician who referred the child for the procedure. | If a referral date cannot be identified, the assign the date of procedure as the index date. |
| 7 | Assign the index encounter date. | For each individual, order all eligible encounters from the one closest to diagnosis to furthest from diagnosis. The encounter furthest from diagnosis is the index date. | Calculate the diagnostic interval as the number of days between the index date and the date of formal diagnosis. |

| Supplementary Table S4. Eligible encounter categories for leukemias | | | | |
|---|---|---|---|---|
| **Category number** | **Encounter type** | **Encounter Category** | **Billing codes** | |
| **Procedure groups** | | | | |
| 1 | Procedure | Anesthesiology | CCI | |
| | | | ACTE | 15487, 15601, 15602, 41001 |
| 2 | Procedure | Biopsy | CCI | 2WY71HA |
| | | | ACTE | 04161, 00212, 04159, 00234, 0273, 0281, 00249, 00282, 00215 |
| 3 | Procedure | Cancer-related encounter | CCI | 1AX35HA, 1ZZ35HA |
| | | | ACTE | 08553, 00595, 08511, 15021, 15005, 15012, 15020, 15007, 15122, 00593, 00596, 15121, 00734, 00094, 09168 |
| 4 | Procedure | Cardiovascular-related encounter | CCI | 1IS53HN, 1KX53HA, 1IS53LA, 1IS53GR |
| | | | ACTE | 00350, 00295, 00751, 08409, 09305, 09333, 20129, 00913, 08401, 09472, 00769, 08360, 00585, 00909, 08668, 09308, 00765, 08303, 09331 |
| 5 | Procedure | Consult or assessment-other speciality | CCI | |
| | | | ACTE | 09283, 16018, 09287, 09288, 09252, 16105, 00043, 09253, 15179, 00036, 09250, 09184, 09255, 08936, 09165, 09162, 09170, 09150, 09160, 08976 |
| 6 | Procedure | Consult or assessment-Paediatrics | CCI | |
| | | | ACTE | 15550, 15186, 15543, 15166, 15548, 15165, 15540 |
| 7 | Procedure | Critical care | CCI | 1GZ31CB, 1GZ31CA |
| | | | ACTE | 00010,15389, 09095, 00928, 15483, 00900, 16060, 09097 |
| 8 | Procedure | CT- chest, abdomen, pelvis | CCI | |
| | | | ACTE | 08263, 08255, 08266, 08268, 08262 |
| 9 | Procedure | CT-head, neck, spine | CCI | |
| | | | ACTE | 08275, 08290, 08260, 08258, 08259 |
| 10 | Procedure | Emergency room | CCI | |
| | | | ACTE | 15598, 15212, 15216, 15461, 15478, 15217, 16020, 09046, 15218, 09108, 15215, 15213, 15210 |
| 11 | Procedure | Hematology-related encounter | CCI | 1LZ20HT, 1LZ35HH, 1LZ19HH |
| | | | ACTE | 20539, 00434 |
| 12 | Procedure | Infectious disease encounter | CCI | |
| | | | ACTE | 09026, 09031, 09027 |
| 13 | Procedure | Medical genetics | CCI | |
| | | | ACTE | 09608, 60062, 60006, 60060, 60002, 60061, 09603, 09660, 09607 |
| 14 | Procedure | Miscellaneous | CCI | 1ZZ35CA, 1FR87LA |
| | | | ACTE | 00127, 08925, 00691, 00519, 00746, 00276, 15101, 20534 |
| 15 | Procedure | MRI-head, neck, spine | CCI | |
| | | | ACTE | 08442, 08443, 08577, 08576, 08578, 08570 |
| 16 | Procedure | Neurology-related encounter | CCI | 2AX13HA |
| | | | ACTE | 00752, 20264, 00347 |
| 17 | Procedure | Other imaging | CCI | |
| | | | ACTE | 08277, 08693, 08280, 08678, 08631, 00627, 20076, 08686, 08639, 08679, 08648, 08243, 08700, 08692, 08703, 08276, 08439 |
| 18 | Procedure | Ultrasound – abdomen, pelvis | CCI | |
| | | | ACTE | 08366, 08325, 08334, 08321, 08399, 08315, 08398, 08326 |
| 19 | Procedure | Ultrasound-other | CCI | |
| | | | ACTE | 08330, 08302, 08331, 08335, 08393, 08342, 08426, 08392, 08346 |
| 20 | Procedure | Xray-chest, abdomen, pelvis | CCI | |
| | | | ACTE | 08100, 08150, 08115, 08152, 08102 |
| 21 | Procedure | Xray- head, neck, spine | CCI | |
| | | | ACTE | 08054, 08042, 08128, 08127, 08059, 08056, 08037, 08125 |

| | | | | |
|---|---|---|---|---|
| 22 | Procedure | Xray- other | CCI | |
| | | | ACTE | 08063, 08062, 08083, 08086, 08065, 08085, 08064, 08060, 08087, 08084, 08080 |
| **Diagnosis groups** | | | | |
| 1 | Diagnostic | Circulatory system diagnoses | ICD-9/10 | 4259, 7850, 4279, 4010, 7943, 4299, 4019, I313, I828, R001, I1590, I959, R000, I100, 7852, I1580, 4289 |
| 2 | Diagnostic | Ear-related diagnoses | ICD-9/10 | 3812, 3811, H669, 3820, 3814, 3829, H659, 3801, 3810, 3887, 3889, 3802, 3823, 3824, 3849, 3886,V721 |
| 3 | Diagnostic | Endocrine and metabolic disorders | ICD-9/10 | 2767, 2779, 2762, 2599 ,7832, 2754, 7906, 250, 2769, 2780, 2789, 2729, 2559, 2535, 7836, 2400, 2449, 3075, 2554, E883, 2512, 2500, E222, E870, E781, E139, E274, E162, E872, E875, R633, E860, E835, E871, E877, R630, R634, R739, R748, E46, E880, R740, E834, E833, E876 |
| 4 | Diagnostic | Infectious disease | ICD-9/10 | 0409, 239, 0419, 1125, R508, 0388, 1109, 3229, 3730, 482, 0091, 7119, 6868, 0380, 0406, 0415, 0790, 1173, 1129, 4879, 0090, 0408, 4609, 4740, 7309, 4210, R509, 1369, 087, 0389, 7908, 0879, 7907, 7806, L0310, T827, T82700, A408, B974, B971, B9688, B962, B957, B3788, T82701, B956, B370, R572, A491, B348, A490, R508, R502 A047, A498, B9788, R509, 465, 480, 4669, 4800, 4859, 4650, J069, 0204, 4829, 4644, 486, 4658, 4660, 4871, 4909, 4809, 4661, 4640, 4629, 4869, 4659, 4618, J189, J069, J00, J101, 0539, 0549 |
| 5 | Diagnostic | Leukemia | ICD-9/10 | 2042, 2060, 2031, C920, 207, 2387, 2058, C959, 2051, 2059, 205, 2029, 2048, 2041, 204, C910, 2049, 2079, C950, 2078, 2050, 2088, 208, 2040, 2089, 2080, C920, C959, C910 |
| 6 | Diagnostic | Other cancer | ICD-9/10 | 2002, 2161, 1719, 2001, 1919, 199, 2169, 1739, 2028, 2399, C809, 1991 |
| 7 | Diagnostic | Respiratory system | ICD-9/10 | 4969, 5119, 7802, 5199, 5198, 7860, 7862, J980, 7489, J9810, J984, R060, R05, R090, 4782, 5188, J90, 7841 |
| 8 | Diagnostic | Routine examination | ICD-9/10 | Z049, Z008, V70, V589, Z001, V708, Z000<br>V709, V202, V700 |
| 9 | Diagnostic | Signs and symptoms – genitourinary | ICD-9/10 | 5908, 7881, 5819, 5901, 5919, 5997, 5849, 5869, 5959, 5990, N390, N179, 5920, 7880, V259, 6069, 5929, 7529 |
| 10 | Diagnostic | Signs and symptoms – GI | ICD-9/10 | 4553, 5278, 5362, 5368, 5559, 5280, 5350, 5645, 5355, 5301, 5649, 7891, 5770, 7870, 5779, 5640, 5589, K625, K521, K729, R160, E806, K9280, K859, K219, R162, K831, K602, R112, K529, A099, R111, R113, K590, K123, 5758, K051, 7833, 5742, 5419, 5693, 5409, 5650, 540, 7872 |
| 11 | Diagnostic | Signs and symptoms – hematology | ICD-9/10 | 2840, 7856, 2872, 2874, 4461, 4519, 6262, 7851, D700, 2898, 2819, 2790, 2848, D729, 2893, 7169, 2875, 2881, 4539, 2860, 2882, 2879, 2799, 7909, 2793, 7847, 2899, 2880, 2849, 2859, 7827, 2889, D65, D695, R798, D689, R040, D649, D696, D630, D619, D700, D728, 7767, 4590, 7769 |
| 12 | Diagnostic | Signs and symptoms – musculoskeletal | ICD-9/10 | R104, 7242, 7291, 7194, 7245, 7295, 7865, 7270, 7330, 7807, 7199, R529, 7239, 7890, M7961, 7822, R53, R104, M7929, 3569, 7569, 7331, 3588, 7110, 7334, 7302, 0781 |
| 13 | Diagnostic | Signs and symptoms – NOS | ICD-9/10 | 7899, 7823, 7580, 7598, 7599, 0759, 7589, 3079, 9959, 7805, 7809, 0799, V498, 9952, V726, V729, T808, 2280, 9950, L270, V263, 7080, R609, R18 |
| 14 | Diagnostic | Signs and symptoms – Vision | ICD-9/10 | 3779, 3688, 379, 7439, 3781, 3792, 3770, 3629, 3689, 3799, V720, H356, 3669, 3780, 366 |
| 15 | Diagnostic | Signs and symptoms – neurology | ICD-9/10 | 7429, 7819, 7803, 3000, 7840, F419, 3469, R51, G629 |

| **Supplementary Table S5.** Eligible encounter categories for lymphomas | | | | |
|---|---|---|---|---|
| **Category number** | **Encounter type** | **Category name** | **Billing codes** | |
| **Procedure groups** | | | | |
| 1 | Procedure | Anesthesiology | CCI | |
| | | | ACTE | 41045, 41049, 41004, 15487, 15602, 15601, 15486 |
| 2 | Procedure | Biopsy – lymph node | CCI | 2MC71HA, 2MJ71LA, 2MD71LA, 1MC87LA, 2MC71LA |
| | | | ACTE | 00252, 00184 |
| 3 | Procedure | Biopsy – other | CCI | 2OA71HA, 2OT71HA, 2GW71HA, 2WY71HA |
| | | | ACTE | 00234, 00281, 00282, 00181, 00202, 00273, 02062, 09466, 00237, 01132, 02175, 02797, 09550, 00212, 00249, 00215, 00308, 09465, 09464 |
| 4 | Procedure | Cancer-related encounter | CCI | 1AX35HA, 1ZZ35HA, 2AX13HA |
| | | | ACTE | 15005, 15007, 15020, 00094, 00734, 00596, 00595, 00593, 15021 |
| 5 | Procedure | Cardiovascular-related encounter | CCI | 1IS53LA, 1IS53GR, 1LZ19HH |
| | | | ACTE | 00294, 00585, 00597, 00639, 08359, 08388, 09436, 00575, 09334, 09308, 00909, 08668, 08360, 00765, 08303, 09472, 09331 |
| 6 | Procedure | Consult or assessment- other specialty | CCI | |
| | | | ACTE | 08937, 09254, 09282, 15101, 16018, 16105, 08976, 09287, 15132, 00042, 09255, 00043, 09253, 09288, 09184, 09250, 00036, 09283, 09165, 09162, 09150, 09160, 08936, 09170 |
| 7 | Procedure | Consult or assessment- paediatrics | CCI | |
| | | | ACTE | 15550, 15543, 15164, 15186, 15166, 15548, 15165, 15540 |
| 8 | Procedure | Critical care | CCI | |
| | | | ACTE | 00940, 00912, 09097, 15483, 09095, 15389 |
| 9 | Procedure | CT-chest, abdomen, pelvis | CCI | |
| | | | ACTE | 08264, 08269, 08256, 08267, 08263, 08268, 08255, 08266, 08262 |
| 10 | Procedure | CT- head, neck, spine | CCI | |
| | | | ACTE | 08261, 08275, 08290, 08259, 08258, 08260 |
| 11 | Procedure | Emergency room | CCI | |
| | | | ACTE | 15215, 15212, 15218, 15216, 09046, 15355, 00051, 15461, 09108, 15213, 15210 |
| 12 | Procedure | Medical genetics | CCI | |
| | | | ACTE | 09607, 09608, 09634, 60060 |
| 13 | Procedure | Miscellaneous | CCI | 1GV52HA |
| | | | ACTE | 00276, 08925, 20184, 08479, 01121, 00127, 09418 |
| 14 | Procedure | MRI- chest, abdomen, pelvis | CCI | |
| | | | ACTE | 08572, 08573, 08574, 08575 |
| 15 | Procedure | MRI-head, neck, spine | CCI | |
| | | | ACTE | 08578, 08576, 08571, 08570 |
| 16 | Procedure | Other imaging | CCI | |
| | | | ACTE | 08686, 08179, 08243, 08703, 08276, 08277, 08693, 08692, 08679, 08700 ,08439, 00627 |
| 17 | Procedure | Surgical resection and exploration | CCI | |
| | | | ACTE | 03035, 03167, 04233, 05010, 07642, 07684, 04160, 01106, 03027, 03195, 05141, 05172, 05480, 03196, 05077, 09362, 09363, 00324, 00697, 04162, 00691, 07525, 00746, 04161, 04159, 05993, 18171, 05201, 05228 |
| 18 | Procedure | Ultrasound – chest, abdomen, pelvis | CCI | |
| | | | ACTE | 08348, 08383, 08334, 08325, 08366, 08321, 08326, 08398, 08399 |
| 19 | Procedure | Ultrasound-other | CCI | |
| | | | ACTE | 08392, 08427, 08343, 08358, 08342, 08331, 08335, 08330 |

| | | | | |
|---|---|---|---|---|
| 20 | Procedure | Xray-chest, abdomen, pelvis | CCI | |
| | | | ACTE | 08115, 08150, 08152, 08100 |
| 21 | Procedure | Xray- head, neck, spine | CCI | |
| | | | ACTE | 08042, 08037, 08058, 08128, 08125 |
| 22 | Procedure | Xray- other | CCI | |
| | | | ACTE | 08080, 08087, 08091, 08060, 08062 |
| **Diagnosis groups** | | | | |
| 1 | Diagnostic | Circulatory system diagnoses | ICD-9/10 | 4010, 7850, 4259, 4239, 4279, 4019, 7943, I959, R000, I100, I313, 4289, 7852, 4299 |
| 2 | Diagnostic | Dermatological conditions | ICD-9/10 | 6961, 6969, 0579, 702, 6959, 6869, 6909, 0780, 6931, 6939, 7089, 6918, 7822, 7099, 7029, 7821, 6929 |
| 3 | Diagnostic | Endocrine and metabolic disorders | ICD-9/10 | 2729, 2769, 2554, 2559, 2779, 7836, 3075, 7832, 2535, 2780, 2789, 2778, 2449, 2500, E860, E877, E871, R748, R630, R739, E46, E880, E833, E834, E876, E883 |
| 4 | Diagnostic | Genito-urinary system | ICD-9/10 | 5901, 5997, 5849, 5869, 5959, 5990, N179, 6069, V259, V25, 6289, V250 |
| 5 | Diagnostic | Infectious diseases | ICD-9/10 | R509, 1369, 0389, 0879, 7908, 7907, 7806, A047, B9788, R509, 486, 4661, 4658, 4809, 4871, 4909, 4640, 4629, 4660, 4869, 4659, 4609, J189, J069, 4740, 0539, 4741 |
| 6 | Diagnostic | Lymphoma | ICD-9/10 | 1960, 2015, 204, 2011, 2014, C837, C817, 202, 2000, C811, C857, C969, 2002, 2022, C819, 2012, 201, 2001, 2010, 2020, 2019, 2029, 2028, C833, C835, C837, C819, C859, C811 |
| 7 | Diagnostic | Musculoskeletal system | ICD-9/10 | 7199, 0781, 7295, 7330, R104, 7807, 7865, 7245, 7242, 7231, 8472, 7890, 7194, 7291, R221, R222, R104 |
| 8 | Diagnostic | Neurological diagnoses | ICD-9/10 | 3459, 3559, 7803, 7805, 0341, 3469, 7840, 3000, M7929, R51, F419 |
| 9 | Diagnostic | Other cancer | ICD-9/10 | 1649, 1709, 1910, 2298, 2078, 2290, 1923, 2088, 1629, 2299, 1999, 2050, 2125, 2396, 1719, 1739, 1919, 2169, 2398, 2089, 2399, 2040, C809, 199, 2080, 1991 |
| 10 | Diagnostic | Respiratory system | ICD-9/10 | 5198, 5199, 5191, 5119, 5192, 5188, 7802, 7489, 7862, 7860, 4720, R91, J984, R090, J90 |
| 11 | Diagnostic | Routine examination | ICD-9/10 | V70, V708, Z001, Z049, Z000, Z008, V202, V709, V700, V589 |
| 12 | Diagnostic | Signs and symptoms – GI | ICD-9/10 | 5770, 5355, 5600, 0529, 7872, 5649, 5693, 7833, 5559, 5645, 5301, 7870, 5640, 5589, R112, A099, R113, K590, R111, K123, 5419 |
| 13 | Diagnostic | Signs and symptoms – hematology | ICD-9/10 | 4519, 2891, 4539, 462, 6839, 2790, 2882, R599, 7909, 6262, 2799, 2880, 7847, 2893, 2023, 2793, 2859, 2899, 2889, C966, D508, I828, D611, D649, D696, D619, R590, D630, D700, 7856 |
| 14 | Diagnostic | Signs and symptoms – NOS | ICD-9/10 | 7823, 4781, 5193, V498, 3079, 3887, 7809, 0799, V726, 7899, V720, 7239, V725, V729, T808, 3781, 7599, 0759, 3629, 7439, 3709, 3689, 3799, 2280, V263, 9952, V428 |

| Supplementary Table S6. Eligible encounter categories for brain and central nervous system tumours | | | | |
|---|---|---|---|---|
| **Category number** | **Code type** | **Category name** | **Billing codes** | |
| **Procedure groups** | | | | |
| 1 | Procedure | Anesthesiology | CCI | |
| | | | ACTE | 15487, 15602,15601, 15486, 41045, 41046, 41004, 41039 |
| 2 | Procedure | Biopsy | CCI | 2AA71SE, 2AC71SZ, 2AE71SZ, 2AW71LA, 2EA71LA, 2AJ71SZ, 2AX71LA, 2BA71SE, 2AE71DA, 2AE71SE, 2AN71SZ, 2AN71SE, 2AP71SZ, 2AC71DA, 2WY71HA |
| | | | ACTE | 07690, 07528, 07531, 07643, 00215, 00282 |
| 3 | Procedure | Cancer-related encounter | CCI | 1ZZ35HA |
| | | | ACTE | 08511, 08504, 08553, 15702, 00734, 15465, 08518, 08519, 08520, 08565, 20158, 09168 |
| 4 | Procedure | Cardiovascular related encounter | CCI | 1LZ35HH, 1KX53HA, 1IS53GR, 1IS53LA, 1LZ19HH |
| | | | ACTE | 0035, 08388, 09307, 08435, 08414, 08668, 08360, 08401, 08585, 00765, 08434, 09308, 08439, 08303 |
| 5 | Procedure | Consult or assessment – other specialty | CCI | |
| | | | ACTE | 00043, 09281, 15344, 15430, 16105, 08937, 09184, 09288, 15346, 00036, 08936, 15333, 09282, 16106, 09254, 09252, 09283, 00080, 09255, 09162, 09150, 09170, 09160, 08976 |
| 6 | Procedure | Consult or assessment – pediatrics | CCI | |
| | | | ACTE | 15186, 15164, 15548, 15166, 15165, 15540, 16100 |
| 7 | Procedure | Critical care | CCI | 1GZ31CA, 1GZ31CB |
| | | | ACTE | 00912, 15389, 15483, 09095, 00928, 00927, 00900, 09097 |
| 8 | Procedure | CT – head, neck, spine | CCI | |
| | | | ACTE | 08275, 08260, 08290, 08258, 08259 |
| 9 | Procedure | Emergency room | CCI | |
| | | | ACTE | 00051, 15218, 15212, 15215, 15216, 15461, 15213, 15210, 09108 |
| 10 | Procedure | Medical genetics | CCI | |
| | | | ACTE | 09601, 09635, 09661, 09634, 09660 |
| 11 | Procedure | Miscellaneous | CCI | |
| | | | ACTE | 05010, 00320, 00691, 08925 |
| 12 | Procedure | MRI – head, neck, spine | CCI | 3ER40VA, 3ER40WC, 3SC40WE, 3SC40VA, 3CA40WE, 3JX40WC, 3JX40VA, 3CA40WA, 3CA40WC, 3SC40WC, 3AN40WA, 3AN40WE, 3AN40WA, 3AN40WC |
| | | | ACTE | 08577, 08571, 08576, 08578, 08570 |
| 13 | Procedure | MRI – other | CCI | |
| | | | ACTE | 08572, 08574, 08442, 08573, 08575 |
| 14 | Procedure | Neurology-related encounter | CCI | 2AN24JA, 2BX25JA, 2AX13HA |
| | | | ACTE | 00357, 00593, 20264, 00145, 00595, 00752, 16070, 00208, 00209, 00347, 07612, 00889, 00596, 00614, 07620 |
| 15 | Procedure | Other brain and CNS procedure | CCI | 1SC80PF, 1SA80PF, 1AC52SZ, 1AC54ME, 1AX52HA, 1SC87PF, 1SC74PF, 2AN28JA, 1AN52SE, 1AC52SE, 1AC55SZ, 1AC52MJ, 1AC52DA, 1AC52ME, 1AC52MB, 1EA80LA, 1AA80SZ |
| | | | ACTE | 07614, 00629 |
| 16 | Procedure | Other imaging | CCI | 3AN94ZA, 3AN94ZB, 3AN94ZC |
| | | | ACTE | 08693, 08634, 08262, 08268, 08277, 08692, 08679, 08700, 20076, 08703 |
| 17 | Procedure | Respiratory-related encounter | CCI | 2GE70BA, 2FA70BA |
| | | | ACTE | 00710, 08133, 00746, 00519 |
| 18 | Procedure | Surgical resection – brain and CNS | CCI | 1AA87SZ, 1AG87SZ, 1AP87SE, 1EA87LA, 1AC87DA, 1AE87SZ, 1AK87SZ, 1AN87SE, 1AW87LA, 1AP87SZ, 1AJ87SZ, 1AN87SZ, 1AC87SZ |
| | | | ACTE | 07524, 07542, 07520, 07521, 07653, 07689, 07534, 07686, 07523, 07529, 07530, 07527, 07652, 07532, 07642, 5962, |

| | | | | |
|---|---|---|---|---|
| | | | | 05956, 05955, 05954, 05961, 05960, 05959, 05958, 05957, 05950, 15724 |
| 19 | Procedure | Ultrasound – head, neck | CCI | |
| | | | ACTE | 08320, 08330, 08302 |
| 20 | Procedure | Ultrasound – other | CCI | |
| | | | ACTE | 08315, 08342, 08366, 08325, 08335, 08321, 08326 |
| 21 | Procedure | Vision-related encounter | CCI | 2CZ02ZZ |
| | | | ACTE | 00553, 00624, 20100, 00860, 20099, 00728, 00579, 15099, 15101, 09125, 09124, 15179 |
| 22 | Procedure | Xray – head, neck, spine | CCI | |
| | | | ACTE | 08037, 08125, 08128, 08042 |
| 23 | Procedure | Xray – other | CCI | |
| | | | ACTE | 08086, 08154, 08152, 08100, 08158 |
| **Diagnosis groups** | | | | |
| 1 | Diagnostic | Brain & CNS tumours | ICD-9/10 | 1915, C716, 2273, Q850, 1922, 2250, 2397, 1929, C717, 2259, 191, 1923, 1916, C719, C710, 1917, D432, 1921, 2396, 1919, 1910, C712, C715, C711, C710, D431, C718, D430, D432, C719, C716, C717 |
| 2 | Diagnostic | Circulatory system | ICD-9/10 | 4309, 4279, 4019, 4590, 7469, 7856, 4299, 7852, 7455, 4539, 7943, I100, R000, R001, G932, 4319 |
| 3 | Diagnostic | Endocrine and metabolic disorders | ICD-9/10 | 2779, 253, 2556, 2593, 2535, 2537, 2780, 7832, 2789, 2534, 2599, 2769, 2500, 3075, 2539, 7834, 6260, 2533, 2559, 2554, 2591, V779, 2532, 2449, E039, E274, E872, R634, R630, E860, E222, R633, E833, E46, E876, E871, E834 |
| 4 | Diagnostic | Signs and symptoms – genitourinary system | ICD-9/10 | 5969, 7883, 5901, 5997, 7882, 5990, 5959, N390 |
| 5 | Diagnostic | Infectious diseases | ICD-9/10 | 3209, 4879, 7908, B956, 3229, 4871, 0389, 0799, 0879, 7806, 87, 0341, R509, R509, 0529, 0549, 0539 |
| 6 | Diagnostic | Neurological disorders | ICD-9/10 | 3009, 3090, 7820, F900, 7800, 3861, F419, 3460, 3483, 438, 462, 4783, 3314, G409, 345, 3489, R51, 3239, 3313, V403, 7429, 3349, 3482, 3119, 3139, 3348, 7804, 3458, 3099, 7423, 3159, 3469, 3000, 3140, 3319, 3459, 7803, 7840, 3480, 3454, 7419, 315, F89, G978, F419, G935, G939, T850, G4090, F900, G510, G911, G919, G9388, G8199, R51, G941, 8509 |
| 7 | Diagnostic | Musculoskeletal diagnoses | ICD-9/10 | 7199, 7339, 3559, 7235, 7819, 7269, 0781, 7295, 7291, 7242, 7194, 7807, 7373 |
| 8 | Diagnostic | Signs and symptoms – nervous system and movement disorders | ICD-9/10 | 344, 3441, 3446, 7812, R5688, R278, R270, 3343, R568, 3154, 3449, 3429, 7813, 7810 |
| 9 | Diagnostic | Other cancer | ICD-9/10 | 2249, 1749, C793, C794, 1983, 2298, 2388, 1890, 2320, 7287, 1739, 2050, 1940, 2299, 2161, 1918, 2089, 2040, 2169, 1999, 2398, 2080, 2252, 2377, C809, 199, 2399, 2381, 1991, C723, Q850, D489 |
| 10 | Diagnostic | Respiratory system | ICD-9/10 | J069, V760, 5199, 7862, 486, 4609, 5191, 4781, 5198, 4809, 4658, 5188, 4660, 4619, 4640, 4909, 4661, 4869, 4779, 4720, 4739, 7860, 7802, 4659, J4500, R068, 4782, J069 |
| 11 | Diagnostic | Routine examination | ICD-9/10 | Z001, Z008, V708, Z000, V589, V709, V202, V700 |
| 12 | Diagnostic | Signs and symptoms – gastrointestinal tract | ICD-9/10 | 5368, 0091, 5362, A047, 0090, 5355, 7872, 5350, 5649, 7833, 7870, 5301, 5640, 5589, K123, R138, A099, R111, R112, K590, R113 |
| 13 | Diagnostic | Signs and symptoms – hematology | ICD-9/10 | D729, 2881, 2882, 2899, 2880, 7909, 2859, 7907, 2889, D611, D649, D619, D630, D696, D700, 7847 |
| 14 | Diagnostic | Signs and symptoms – NOS | ICD-9/10 | 7069, 7823, 7599, 3079, 7805, V498, 7809, V726, 7865, 7245, V725, V729, 7890, 6909, R104, 7589, V441, V549, V829, V263 |
| 15 | Diagnostic | Signs and symptoms - vision | ICD-9/10 | 3682, 3743, 377, 3659, 3669, 7439, 3629, 3771, 3795, 3781, 3780, 3770, 3689, 3789, 3779, 379, 3799, V720, H500, H534, H55, H532, H492, H471 |

| | | | | |
|---|---|---|---|---|
| 16 | Diagnostic | Signs and symptoms – speech, language, hearing | ICD-9/10 | 3153, 3158, 3811, 3820, 3887, 3814, H669, 3829, 3802, 3899, 3810, 3889, 3801, 3897, 3804, 7844, 7845, H910, H919, R470 |

**Supplementary Table S7.** Eligible encounter categories for neuroblastoma and peripheral nervous system tumours

| Category number | Code type | Category name | | Billing codes |
|---|---|---|---|---|
| **Procedure groups** | | | | |
| 1 | Procedure | Anesthesiology | CCI | |
| | | | ACTE | 15487, 15602, 41039, 41050, 41049, 41001, 41004, 41045 |
| 2 | Procedure | Biopsy | CCI | 2GW71DA, 2OA71LA, 2OT71HA, 2SZ71LA, 2OT71LA, 2WY71HA, 2MC71LA, 2MG71DA, 2MG71LA |
| | | | ACTE | 00281, 09465, 00273, 00249, 00282 |
| 3 | Procedure | Cancer-related encounter | CCI | 1AX35HH, 1ZZ35HA |
| | | | ACTE | 00596, 00595, 00734, 15007, 15021, 09168 |
| 4 | Procedure | Cardiovascular-related encounter | CCI | 1IS53HN, 1IS53GR, 1IS53LA, 1LZ19HH, 1LZ35HH |
| | | | ACTE | 08388, 08292, 00765, 08360, 08668, 09308, 08303, 00769, 08061 |
| 5 | Procedure | Consult or assessment – other speciality | CCI | |
| | | | ACTE | 00036, 09162, 09170, 09150, 09160, 09127, 09250, 09283, 09165, 15389, 16105, 09255, 08936, 08976 |
| 6 | Procedure | Critical care | CCI | 1GZ31CA, 1GZ31CB |
| | | | ACTE | 00912, 00928, 00940, 09097 |
| 7 | Procedure | CT – chest, abdomen, pelvis | CCI | |
| | | | ACTE | 08263, 08264, 08266, 08255, 08262, 08268 |
| 8 | Procedure | CT – head, neck, spine | CCI | |
| | | | ACTE | 08290, 08259, 08258, 08260 |
| 9 | Procedure | Emergency room | CCI | |
| | | | ACTE | 15213, 15210, 09108, 15598 |
| 10 | Procedure | Miscellaneous | CCI | |
| | | | ACTE | 09634, 09660, 04233, 00127, 09472, 00357, 08925 |
| 11 | Procedure | Other imaging | CCI | |
| | | | ACTE | 08693, 00363, 08679, 08700, 08692, 08575, 08703 |
| 12 | Procedure | MRI – chest, abdomen, pelvis | CCI | 3OT40WC, 3OT40WA |
| | | | ACTE | 08574, 08572, 08573 |
| 13 | Procedure | MRI – head, neck, spine | CCI | 3AN40WA, 3AN40WC |
| | | | ACTE | 08571, 08578, 08570, 08577, 08576 |
| 14 | Procedure | Consult or assessment - pediatrics | CCI | |
| | | | ACTE | 15166, 15548, 15186, 15165, 15540 |
| 15 | Procedure | Surgical resection of thorax, abdominal, and retroperitoneal structures | CCI | 1EQ87LA, 1SZ87LA, 1GW87LA, 1PB87LB, 1GW87DA, 1OT91LA, 1PB89DA, 1PB89LB, 2OT70LA |
| | | | ACTE | 05145, 06182, 03196, 05077, 05480, 04159, 04161, 05938, 05939, 03011 |
| 16 | Procedure | Ultrasound – chest, abdomen, pelvis | CCI | |
| | | | ACTE | 08331, 08398, 08325, 08399, 08366, 08326, 08335 |
| 17 | Procedure | Genitourinary-related encounters | CCI | |
| | | | ACTE | 08634, 08639, 08690, 08611, 08635, 08637, 00372 |
| 18 | Procedure | Xray – chest, abdomen, pelvis | CCI | |
| | | | ACTE | 08150, 08152, 08102, 08100, 08055 |
| **Diagnosis groups** | | | | |
| 1 | Diagnoses | Circulatory system | ICD-9/10 | 4059, 7856 |
| 2 | Diagnoses | Endocrine and metabolic disorders | ICD-9/10 | 2558, 2599, 2449, 255, 2554, 259, 2500, 2512, 3075, 2762, 2768, 2769, V779, 2559, E872, R633, E875, E871, E46, E833, E834, E876, E835, R748, R739, 7832, R634, E860, R630, |

| | | | | |
|---|---|---|---|---|
| | | | | R740, 7831 |
| 3 | Diagnoses | Signs and symptoms – genitourinary system | ICD-9/10 | N133, R311, R33, N179, 5965, 7539, 7883, 5969, 7882, 5997, 5819, V819, 5919, 5849 |
| 4 | Diagnoses | Infectious disease | ICD-9/10 | 0415, 6869, 0790, 4619, R509, 0799, 7806, R509, R508, 7309, 4609 |
| 5 | Diagnoses | Mental health and neurological diagnoses | ICD-9/10 | 3000, 3332, 3339, 3348, 349, 3349, 7429, 7819 7812, 7813, 3588, 7840, 3559, M7929, 3441, 3449 |
| 6 | Diagnoses | Musculoskeletal system | ICD-9/10 | 7119, 7169, 7291, 7194, 7235, 7270, 7373, 7295, 7199, R53, M7961 |
| 7 | Diagnoses | Neuroblastoma and other PNS tumours | ICD-9/10 | 1929, 1948, 2372, 2270, C749, 1949, 192, 2377, 194, 1940, C473, C474, C479, C480, C749 |
| 8 | Diagnoses | Other cancer | ICD-9/10 | 1570, 1899, 1912, 1922, 2000, 2125, 2299, 1987, 2320, 2381, C950, 1952, 1962, 2012, 2130, C719, C761, V169, 1719, 1921, 2155, 1588, 1739, 2029, 1580, 2169, 1589, 1709, 2395, 1890, 2390, C762, 1999, Q850, 2398, 2050, C809, 1928, 2040, 2089, 1919, 2399, 1910, 199, 1923, 2080, 2396, 1600, 1991, C762, C770, C382, C771, C778, C787, C772, C795 |
| 9 | Diagnoses | Respiratory system | ICD-9/10 | 4969, 5119, R05, 7489, 7861, 7860, 7862, J9609, R061, R068, J9810, R060, J90 |
| 10 | Diagnoses | Routine examination | ICD-9/10 | Z001, Z008, V708, Z000, Z049, V909, V589, V709, V202, V700, V209 |
| 11 | Diagnoses | Signs and symptoms – gastrointestinal tract | ICD-9/10 | 7519, 7893, 5608, 5609, 5649, 5640, R111, R112, K590, 7870, R113, 7872, 7833 |
| 12 | Diagnoses | Signs and symptoms – hematological conditions | ICD-9/10 | 2875, 2879, D729, 2893, 7909, 2899, 2859, 2889, R798, D728, D695, D696 |
| 13 | Diagnoses | Signs and symptoms – NOS | ICD-9/10 | R21, 3079, 7964, R104, 7891, R190, 7809, V498, 7807, 7865, 7899, 7890, R590, R222, R18, R190, R529, R104 |
| 14 | Diagnoses | Signs and symptoms - vision | ICD-9/10 | 3799, 3688, 3795, 3780, 3789, 379, 3781, H570, G902 |

**Supplementary Table S8.** Eligible encounter categories for retinoblastoma

| Category number | Code type | Category name | Billing codes | |
|---|---|---|---|---|
| **Procedure groups** | | | | |
| 1 | Procedure | Biopsy | CCI | 2WY71HA |
| | | | ACTE | 00249, 00273, 00282 |
| 2 | Procedure | Cancer-related encounter | CCI | 1ZZ35HA, |
| | | | ACTE | 00595, 15535, 15532, 00734, 00596, 15533, 09168 |
| 3 | Procedure | Cardiovascular-related encounter | CCI | 1IS53GR, 1IS53LA |
| | | | ACTE | 08340, 09436, 08401, 00765, 09308 |
| 4 | Procedure | Consult or assessment – other specialty | CCI | |
| | | | ACTE | 09250, 09254, 09150, 09160, 09255, 09162, 09283, 09170, 15485 |
| 5 | Procedure | Consult or assessment - pediatrics | CCI | |
| | | | ACTE | 15186, 15548, 15165, 15540 |
| 6 | Procedure | CT – head, neck, spine | CCI | |
| | | | ACTE | 08259, 08258, 08290 |
| 7 | Procedure | Emergency room | CCI | |
| | | | ACTE | 15182, 15210 |
| 8 | Procedure | Miscellaneous | CCI | 2AX13HA |
| | | | ACTE | 08925 |
| 9 | Procedure | MRI – head | CCI | |
| | | | ACTE | 08570 |
| 10 | Procedure | Ophthalmology | CCI | 2CZ70JA, 2CZ28JA, 2CZ02ZZ |
| | | | ACTE | 15099, 15101, 00579, 20155, 00860 |
| 11 | Procedure | Surgical resection of ocular structures | CCI | 1CN59LA, 1CP89LA |
| | | | ACTE | 01355, 07356, 07403, 07314, 07133, 07311, 07134 |
| | | | CCI | |

| | | | | |
|---|---|---|---|---|
| 12 | Procedure | Ultrasound – other | ACTE | 08335, 08320 |
| **Diagnosis groups** | | | | |
| 1 | Diagnoses | Infectious disease | ICD-9/10 | 0879, R509, 7908, 7806 |
| 2 | Diagnoses | Mental health and neurological diagnoses | ICD-9/10 | 0781, 3159, 8540 |
| 3 | Diagnoses | Other cancers | ICD-9/10 | 1739, 1919, C809, 2080, 199, 1991 |
| 4 | Diagnoses | Retinoblastoma | ICD-9/10 | 1908, 190, C699, 1909, 1905, C692, Z031 |
| 5 | Diagnoses | Routine examination | ICD-9/10 | V708, V726, V909, V709, V729, V202, V700, V263 |
| 6 | Diagnoses | Signs and symptoms – GI | ICD-9/10 | 7870, R112, R111, R113 |
| 7 | Diagnoses | Signs and symptoms – NOS | ICD-9/10 | R609, 6820 |
| 8 | Diagnoses | Signs and symptoms – vision | ICD-9/10 | 3769, 3780, 3622, 3789, 379, 7439, 3621, 3799, 3688, 3629, V720, 3689, 3723 |

**Supplementary Table S9.** Eligible encounter categories for renal tumours

| Category number | Code type | Category name | Billing codes | |
|---|---|---|---|---|
| **Procedure groups** | | | | |
| 1 | Procedure | Anesthesiology | CCI | |
| | | | ACTE | 00297, 41004, 41039, 41001, 41050, 41049 |
| 2 | Procedure | Biopsy | CCI | 2PC71LA, 2PC71HA |
| | | | ACTE | 00252 |
| 3 | Procedure | Biopsy – other | CCI | 2ME71LA, 2MH71LA, 2WY71HA, 1MH87LA, 2MG71LA, 1MG87LA |
| | | | ACTE | 09465, 00282 |
| 4 | Procedure | Cancer-related encounter | CCI | 1LZ35HH, 1ZZ35HA, 1AX35HH |
| | | | ACTE | 15020, 00734 |
| 5 | Procedure | Cardiovascular-related encounter | CCI | 1IS53GR, 1IS53LA, 1LZ19HH |
| | | | ACTE | 00585, 00295, 08398, 00765, 08360, 08366, 09308, 09168 |
| 6 | Procedure | Consult or assessment – other specialty | CCI | |
| | | | ACTE | 15487, 15602, 08976, 09221, 16105, 00080, 08936, 09194, 09165, 09162, 09170, 09150, 09160 |
| 7 | Procedure | Consult or assessment – pediatrics | CCI | |
| | | | ACTE | 15186, 15548, 15541, 15165, 15540 |
| 8 | Procedure | Critical care | CCI | 1GZ31CA, 1GZ31CB |
| | | | ACTE | 09097, 15389, 00900 |
| 9 | Procedure | CT – chest, abdomen, pelvis | CCI | |
| | | | ACTE | 08255, 08266, 08262, 08268 |
| 10 | Procedure | Emergency room | CCI | |
| | | | ACTE | 60702, 15215, 15213, 15598 |
| 11 | Procedure | Genito-urinary-related encounter | CCI | |
| | | | ACTE | 00127, 08635, 08637, 08639, 08690, 08634 |
| 12 | Procedure | Miscellaneous | CCI | |
| | | | ACTE | 09660, 08925 |
| 13 | Procedure | MRI – chest, abdomen, pelvis | CCI | |
| | | | ACTE | 08573 |
| 14 | Procedure | MRI – head, neck, spine | CCI | |
| | | | ACTE | 08570, 08578 |
| 15 | Procedure | Other imaging | CCI | |
| | | | ACTE | 08700, 08703, 08692 |
| 16 | Procedure | Surgical resection of renal tissue | CCI | 1PC91QF, 1MH89LA, 1PC89LB, 1PC87LA, 1PC91PF, 1PC91LB |
| | | | ACTE | 06084, 06199, 05480, 06101, 06200, 05942, 05969, 05973, 05938, 05941, 05947, 05972, 05978, 05937, 05945, 05970, 05971, 05977, 05946, 05939, 05940 |
| | | | CCI | |

| | | | | |
|---|---|---|---|---|
| 17 | Procedure | Ultrasound | ACTE | 08359, 08335, 08325, 08399, 08321, 08326 |
| 18 | Procedure | Xray – chest, abdomen, pelvis | CCI | |
| | | | ACTE | 08150, 08100, 08102 |
| **Diagnosis groups** | | | | |
| 1 | Diagnoses | Circulatory system | ICD-9/10 | 4059, 4010, 4019, 7469, R000, I1590, I1580, I1510, I100 |
| 2 | Diagnoses | Endocrine and metabolic disorders | ICD-9/10 | V779, E833, E877, E46 |
| 3 | Diagnoses | Infectious diseases | ICD-9/10 | 0879, 7806, R509 |
| 4 | Diagnoses | Other cancer | ICD-9/10 | 1999, 1910, 1919, 1719, 2089, 1923, 2080, C809, 2399, 199, 1991, C7809, C780, C7801, C772, C7800 |
| 5 | Diagnoses | Renal tumours | ICD-9/10 | 1952, 2369, 1898, 2230, C64, 2395, 1899, 1890, 189, C64 |
| 6 | Diagnoses | Respiratory system | ICD-9/10 | 7860, J9810, J984, J90 |
| 7 | Diagnoses | Routine encounters | ICD-9/10 | V709, V202, V700 |
| 8 | Diagnoses | Signs and symptoms – Genito-urinary system | ICD-9/10 | 5849, 588, 5908, 5919, 5932, 5869, 5819, 5839, 7533, 5938, 5997, 5990, N179, N289, N390, R310, N2888 |
| 9 | Diagnoses | Signs and symptoms – GI | ICD-9/10 | 7870, K566, K565, R111, R113, R112 |
| 10 | Diagnoses | Signs and symptoms – hematology | ICD-9/10 | 2859, 2889, R798, D649 |
| 11 | Diagnoses | Signs and symptoms – NOS | ICD-9/10 | 7809, 7822, 7893, V584, V725, V726, V263, 7599, V769, V729, 7899, R222, R104, R190, 7589, 7890 |

**Supplementary Table S10.** Eligible encounter categories for hepatic tumours

| Category number | Code type | Category name | Billing codes | |
|---|---|---|---|---|
| **Procedure groups** | | | | |
| 1 | Procedure | Biopsy | CCI | 2OA71LA, 2OA71HA |
| | | | ACTE | 00181, 05077, 09465, 09466, 00282 |
| 2 | Procedure | Cancer-related encounter | CCI | 1ZZ35HA |
| | | | ACTE | 15005, 00734, 09168 |
| 3 | Procedure | Cardiovascular-related encounter | CCI | 1LZ35HH, 1IS53LA, 1IS53GR, 1LZ19HH |
| | | | ACTE | 08668, 09221, 09308, 09436, 08401, 00765 |
| 4 | Procedure | Consult or assessment – other specialty | CCI | |
| | | | ACTE | 15210, 09060, 09183, 09250, 00080, 09184, 15389, 09162, 09170, 09150, 09160, 15602, 15485 |
| 5 | Procedure | Consult or assessment. – pediatrics | CCI | |
| | | | ACTE | 15166, 15165, 15540, 15543 |
| 6 | Procedure | CT | CCI | |
| | | | ACTE | 08438, 08263, 08703, 08262, 08268, 08255, 08266 |
| 7 | Procedure | MRI | CCI | |
| | | | ACTE | 08573 |
| 8 | Procedure | Other imaging | CCI | |
| | | | ACTE | 08679, 08692, 08700, 08639 |
| 9 | Procedure | Miscellaneous | CCI | |
| | | | ACTE | 00691, 41049, 08925 |
| 10 | Procedure | Exploration and surgical resection – hepatic tissue | CCI | 1OA87LA |
| | | | ACTE | 05146, 05295, 05939 |
| 11 | Procedure | Ultrasound – chest, abdomen, pelvis | CCI | |
| | | | ACTE | 08366, 08398, 08325, 08335, 08326 |
| 12 | Procedure | Xray – chest, abdomen, pelvis | CCI | |
| | | | ACTE | 08152 |
| **Diagnosis groups** | | | | |
| 1 | Diagnoses | Circulatory system | ICD-9/10 | 4019, 7469, I1590, I1580, R000 |
| 2 | Diagnoses | Endocrine and metabolic disorders | ICD-9/10 | E274, E877, E880, E871, E872, E46, E875, R740, E833, E876, E834 |
| 3 | Diagnoses | Hepatic tumours | ICD-9/10 | C229, 155, 1559, 1550, 1552, C220, C222, Z76804, Z944 |
| 4 | Diagnoses | Infectious disease | ICD-9/10 | 7907, 0879, 5733, 7806, B9788, T814, R509 |
| 5 | Diagnoses | Other cancer | ICD-9/10 | 2399, 199, 1991, C7800, C7801 |

| | | | | |
|---|---|---|---|---|
| 6 | Diagnoses | Respiratory system | ICD-9/10 | 7860, 4869, 7862, J984 |
| 7 | Diagnoses | Routine encounter | ICD-9/10 | V202, V700 |
| 8 | Diagnoses | Signs and symptoms – GI | ICD-9/10 | 7519, 7870, R113, R111 |
| 9 | Diagnoses | Signs and symptoms – hepatology | ICD-9/10 | 5738, 7891, 5739, K769, R104, K831, K729, R190 |
| 10 | Diagnoses | Signs and symptoms – haematology | ICD-9/10 | 4539, 4590, D689, D649 |
| 11 | Diagnoses | Signs and symptoms – NOS | ICD-9/10 | 7899, V729, 7890, N179 |

**Supplementary Table S11.** Eligible encounter categories for malignant bone tumours

| Category number | Code type | Category name | Billing codes | |
|---|---|---|---|---|
| **Procedure groups** | | | | |
| 1 | Procedure | Anesthesiology | CCI | |
| | | | ACTE | 41045, 41039, 41046, 41050, 41049, 15486, 15602, 15601, 15487, 15485, 15600, 70000 |
| 2 | Procedure | Biopsy | CCI | 2SH71HA, 2SQ71HA, 2SQ71LA, 2SW71HA, 2TK71LA, 2SC71HA, 2VQ71HA, 2VC71HA, 2VQ71LA, 2WY71HA, 2VC71LA |
| | | | ACTE | 00249, 00273, 02062, 02084, 02109, 02939, 02992, 09465, 02175, 02796, 09464, 00281, 02864, 00202, 00282, 02865, 00212, 02797 |
| 3 | Procedure | Cancer-related encounter | CCI | 1AX35HH, 1LZ35HH, 1ZZ35HA |
| | | | ACTE | 15131, 15007, 15121, 15021, 15020, 09168, 00734 |
| 4 | Procedure | Cardiovascular-related encounter | CCI | 1IS53GR, 1LZ19HH, 1IS53LA |
| | | | ACTE | 08659, 08670, 09333, 08303, 09308 |
| 5 | Procedure | Consult or assessment – other specialty | CCI | |
| | | | ACTE | 00043, 08936, 09288, 08986, 09165, 15132, 09150, 09162, 09160, 09170, 09097 |
| 6 | Procedure | Consult or assessment – pediatrics | CCI | |
| | | | ACTE | 15186, 15165, 15548, 15540 |
| 7 | Procedure | CT – chest, abdomen, pelvis | CCI | |
| | | | ACTE | 08267, 08268, 08255, 08266, 08263, 08262 |
| 8 | Procedure | CT – head, neck, spine | CCI | |
| | | | ACTE | 08274, 08260, 08290, 08259, 08258, 08275 |
| 9 | Procedure | Emergency room | CCI | |
| | | | ACTE | 09108, 15213, 15215, 60702, 15210 |
| 10 | Procedure | Miscellaneous | CCI | |
| | | | ACTE | 08639, 09607, 08925, 06958 |
| 11 | Procedure | MRI – head, neck, spine | CCI | |
| | | | ACTE | 08573, 08574, 08577, 08576, 08578, 08570 |
| 12 | Procedure | Other imaging | CCI | 3AN94ZB |
| | | | ACTE | 08693, 08692, 08700, 08679, 08276, 08277, 08703, 08575 |
| 13 | Procedure | Respiratory-related encounter | CCI | 1GV52HA |
| | | | ACTE | 09418, 00912, 03122, 00940 |
| 14 | Procedure | Surgical exploration and resection | CCI | 1GR87DA, 1TK87LA, 1SC80PF, 1VC87LA, 1VQ87LA |
| | | | ACTE | 01134, 01205, 02082, 02153, 05480, 07684, 05900, 05938, 05997, 02188, 09567, 18173, 18171, 18172 |
| 15 | Procedure | Ultrasound | CCI | |
| | | | ACTE | 08333, 08325, 08342, 08366, 08360, 08331, 08346, 08392, 08321, 08326, 08335 |
| 16 | Procedure | Xray – extremities | CCI | |
| | | | ACTE | 08062, 08080, 08085, 08083, 08084 |
| 17 | Procedure | Xray – other | CCI | |
| | | | ACTE | 08056, 08102, 08042, 08053, 08054, 08127, 08059 |
| **Diagnosis groups** | | | | |

| | | | | |
|---|---|---|---|---|
| 1 | Diagnoses | Bone tumours | ICD-9/10 | 1704, 1708, C402, 2139, 2392, C419, 1707, 1709, C413, D480, C409, C412, C4100, C414, C400, C419, C402 |
| 2 | Diagnoses | Endocrine and metabolic disorders | ICD-9/10 | E835, R634, E860, E875, E871, E877, R740, R630, R748, E833, E834, E876 |
| 3 | Diagnoses | Musculoskeletal diagnoses | ICD-9/10 | 7179, 8230, 7194, 8210, 7291, 7302, 8290, 9249, 7245, 8449, V549, 0781, 9599, 7199, 7295, R222, L0311, M7961, M907, M7929 |
| 4 | Diagnoses | Other cancer | ICD-9/10 | 1910, 1890, 1999, 1629, 1739, 2169, 1985, 1919, C809, C461, 1923, 2399, 199, 1719, 1991, C780, C782, C7988, C7809, C795, C7801, C7800 |
| 5 | Diagnoses | Respiratory system | ICD-9/10 | 5188, 4869, 7862, 7860, R05, R060, R090, R91, J90 |
| 6 | Diagnoses | Routine encounters | ICD-9/10 | V709, V202, V729, V700 |
| 7 | Diagnoses | Signs and symptoms – urinary system | ICD-9/10 | 7882, 5990, N390 |
| 8 | Diagnoses | Signs and symptoms – GI | ICD-9/10 | 7870, 5640, R112, K123, R113, R111, K590 |
| 9 | Diagnoses | Signs and symptoms – hematology | ICD-9/10 | 2859, D695, T810, R040, R798, D649, D696, D700 |
| 10 | Diagnoses | Signs and symptoms – NOS | ICD-9/10 | 7822, 7899, 7589, V725, 7865, 7890, 7599, R53, 7807, R529, R104, 6260, 6289 |

**Supplementary Table S12.** Eligible encounter categories for sarcomas

| Category number | Code type | Category name | Billing codes | |
|---|---|---|---|---|
| **Procedure groups** | | | | |
| 1 | Procedure | Anesthesiology | CCI | |
| | | | ACTE | 41045, 41046, 41049, 41004 |
| 2 | Procedure | Biopsy | CCI | 2EQ71LA, 2OT71HA, 2SZ71HA, 2EQ71HA, 2SZ71LA, 2VX71LA, 2WY71HA |
| | | | ACTE | 00215, 00273, 09464, 00184, 00281, 01134, 02127, 09465, 00202, 00282 |
| 3 | Procedure | Cancer-related encounters | CCI | 1AX35HH, 1LZ35HH, 1ZZ35HA |
| | | | ACTE | 08504, 15020, 00734, 15021, 20158, 00596, 08511, 08520, 08553, 15465, 09168, 08518, 08519, 08565 |
| 4 | Procedure | Cardiovascular-related encounters | CCI | 1LZ19HH, 1IS53GR, 1IS53LA |
| | | | ACTE | 00295, 00909, 00769, 00765, 08668, 09308, 08360 |
| 5 | Procedure | Consult or assessment – other specialty | CCI | |
| | | | ACTE | 15602, 09149, 09288, 15101, 15461, 15487, 09253, 09255, 09287, 15132, 09250, 09283, 09165, 09127, 09162, 09150, 09160, 09170 |
| 6 | Procedure | Consult or assessment – pediatrics | CCI | |
| | | | ACTE | 15543, 15166, 15186, 15548, 15165, 15540 |
| 7 | Procedure | Critical care | CCI | |
| | | | ACTE | 00900, 00940, 15389, 09097 |
| 8 | Procedure | CT – chest, abdomen, pelvis | CCI | |
| | | | ACTE | 08269, 08267, 08263, 08255, 08266, 08268, 08262 |
| 9 | Procedure | CT – head, neck, spine | CCI | |
| | | | ACTE | 08290, 08260, 08259, 08258 |
| 10 | Procedure | CT – other | CCI | |
| | | | ACTE | 08276, 08277, 08703 |
| 11 | Procedure | Emergency room | CCI | |
| | | | ACTE | 15598, 09108, 15210 |
| 12 | Procedure | Exploration and surgical resection – abdomen | CCI | 1OT87LA, 1YS87LA, 2OT70DA, 1SZ87LA, 1OT52HA, 1GV52HA |
| | | | ACTE | 05010, 05077, 05011, 05480, 09472, 09566 |
| | | | CCI | 1QM91LB, 1WV87LA, 1EQ87LA, 1VX87LA |

| | | | | |
|---|---|---|---|---|
| 13 | Procedure | Exploration and surgical resection – other | ACTE | 06062, 04161, 07642, 05937, 06125, 06309, 02152, 01121, 01205, 04159, 07525, 02153, 18171, 00519, 03209, 09418, 00746 |
| 14 | Procedure | Genitourinary-related encounter | CCI | 1PM52HH, 2PM70BA |
| | | | ACTE | 00127, 00320 |
| 15 | Procedure | Miscellaneous | CCI | |
| | | | ACTE | 09607, 00579, 08925 |
| 16 | Procedure | MRI – abdomen, pelvis | CCI | |
| | | | ACTE | 08573, 08572, 08574 |
| 17 | Procedure | MRI – head, neck, spine | CCI | 3AN40WA, 3AN40WC |
| | | | ACTE | 08578, 08571, 08570 |
| 18 | Procedure | Other imaging | CCI | 3AN94ZB |
| | | | ACTE | 20184, 08692, 08679, 08700, 08575 |
| 19 | Procedure | Ultrasound – other | CCI | |
| | | | ACTE | 08333, 08331, 08392, 08342, 08366, 08325, 08330, 08334, 08303, 08335, 08321, 08326 |
| 20 | Procedure | Xray – chest, abdomen, pelvis | CCI | |
| | | | ACTE | 08115, 08152, 08100 |
| 21 | Procedure | Xray – other | CCI | |
| | | | ACTE | 08056, 08067, 08083, 08084 |
| **Diagnosis groups** | | | | |
| 1 | Diagnoses | Circulatory system | ICD-9/10 | 4019, 7856, 7943, I100 |
| 2 | Diagnoses | Endocrine and metabolic disorders | ICD-9/10 | R739, E860, E46, E834, E833, E876, R630 |
| 3 | Diagnoses | Infectious diseases | ICD-9/10 | R509, 0879, 7806, R509 |
| 4 | Diagnoses | Musculoskeletal diagnoses | ICD-9/10 | 7291, 7245, 7295, 7199, 7865, 7890, R190, R220, R529, R104, R222, 7822 |
| 5 | Diagnoses | Other cancer | ICD-9/10 | 1869, 2019, 2089, 2299, 2396, 1910, 1950, 2392, 2169, 2000, 2398, C809, 2080, 1739, 2053, 1919, 2399, 2012, 199, 1991, C793, C7801, C786, C7800, C795 |
| 6 | Diagnoses | Respiratory system | ICD-9/10 | 4640, 4869, 7862, 7860, J9810, J90 |
| 7 | Diagnoses | Routine examination | ICD-9/10 | V709, V202, V700 |
| 8 | Diagnoses | Signs and symptoms – ear | ICD-9/10 | 3802, 3810, 3801, 3829 |
| 9 | Diagnoses | Signs and symptoms – genitourinary system | ICD-9/10 | 7882, 5919, 5990, R33, N179, N133, N390 |
| 10 | Diagnoses | signs and symptoms – GI | ICD-9/10 | 5640, 7870, 5589, A099, R112, R111, K590, R113 |
| 11 | Diagnoses | Signs and symptoms – haematology | ICD-9/10 | 2859, 2889, D611, D619, D649, D696, D630, D700, T810 |
| 12 | Diagnoses | Signs and symptoms – NOS | ICD-9/10 | 7029, V829, 7062, V726, 7840, 7803, V725, 7899, 7599, V729, V723, 8798, 6820, 6829, 3799, V720 |
| 13 | Diagnoses | Soft tissue and other extraosseous sarcomas | ICD-9/10 | 2381, 1923, C461, 1709, 1719, C491, C494, C495, C490, C492, C499 |

**Supplementary Table S13.** Eligible encounter categories for germ cell tumours, trophoblastic tumours, and neoplasms of gonads

| Category number | Code type | Category name | Billing codes | |
|---|---|---|---|---|
| **Procedure groups** | | | | |
| 1 | Procedure | Anesthesiology | CCI | |
| | | | ACTE | 41001, 41039, 41046, 00255, 41004, 41045, 41049 |
| 2 | Procedure | Biopsy | CCI | 2AF71QS, 2MG71LA, 2WY71HA, 2MH71LA, 2AG71DA, 2AC71DA, 2OT71LA |
| | | | ACTE | 00202 |
| 3 | Procedure | Cancer-related encounter | CCI | 1LZ35HH, 1AX35HH, 1ZZ35HA |
| | | | ACTE | 00734, 15143, 09168 |
| | | | CCI | 1IS53LA |

| | | | | |
|---|---|---|---|---|
| 4 | Procedure | Cardiovascular-related encounter | ACTE | 08401, 08435, 00301, 00765, 09308, 08360, 08303 |
| 5 | Procedure | Consult or assessment – other specialty | CCI | |
| | | | ACTE | 08787, 08937, 09125, 09151, 09287, 15101, 16105, 16106, 00036, 00080, 00579, 08936, 09194, 09288, 09175, 09283, 15389, 09255, 09165, 09162, 09150, 09160, 09170, 15486, 15487, 15601, 15602, 15724, 08976 |
| 6 | Procedure | Consult or assessment – pediatrics | CCI | |
| | | | ACTE | 15543, 15186, 15548, 15165, 15540 |
| 7 | Procedure | CT – head, neck, spine | CCI | |
| | | | ACTE | 08261, 08275, 08290, 08260, 08258, 08259 |
| 8 | Procedure | CT – chest, abdomen, pelvis | CCI | |
| | | | ACTE | 08263, 08262, 08255, 08266, 08268 |
| 9 | Procedure | Emergency room | CCI | |
| | | | ACTE | 15212, 15213, 15215, 15217, 15218, 09108, 15598 |
| 10 | Procedure | Exploration and surgical resection | CCI | 1MG87LA, 1SH87LA, 2OT70LA, 1AC52MB, 1MH87LA, 1OT87DA, 1AC52DA, 2OT70DA, 1OT87LA |
| | | | ACTE | 00614, 00691, 05077, 05954, 07652, 05480, 07642, 07620 |
| 11 | Procedure | Exploration and surgical resection – sex organs | CCI | 1RB87DA, 1RB89LA, 1RD89DA, 1QM89LA, 1RB87LA, 1QM89WJ, 1RD89LA, 1QM91LB |
| | | | ACTE | 06177, 06260, 06188, 06261, 06191, 06309, 06125 |
| 12 | Procedure | Miscellaneous | CCI | 1LZ19HH, 2AX13HA |
| | | | ACTE | 06958, 00596, 08925 |
| 13 | Procedure | MRI – abdomen, pelvis | CCI | |
| | | | ACTE | 08573, 08574, 08578, 08570 |
| 14 | Procedure | Other imaging | CCI | 3AN94ZB |
| | | | ACTE | 08668, 08690, 08692, 09477, 08639, 08679, 08575, 08700, 08703 |
| 15 | Procedure | Ultrasound | CCI | |
| | | | ACTE | 08325, 08330, 08335, 08334, 08321, 08326 |
| 16 | Procedure | Xray | CCI | |
| | | | ACTE | 08150, 08152, 08100 |
| **Diagnosis groups** | | | | |
| 1 | Diagnoses | Endocrine and metabolic disorders | ICD-9/10 | 2535, 2500, 2539, 2449, 2532, E860, E870, E871, E875, E230, E274, E232, E833 |
| 2 | Diagnoses | Germ cell tumours | ICD-9/10 | 2209, C629, C639, 1921, 183, 1879, 186, 2395, 2396, 1919, 1830, 1910, 1869, C719, D27, C6210, C6211, C6291, C6290, C560 |
| 3 | Diagnoses | Infectious diseases | ICD-9/10 | 7907, 7806, R509 |
| 4 | Diagnoses | Musculoskeletal diagnoses | ICD-9/10 | 7245, 7199, 0781, 7807, 7295 |
| 5 | Diagnoses | Mental health and neurological diagnoses | ICD-9/10 | 7803, 3459, 8540, 7840, G941, 3000 |
| 6 | Diagnoses | Other abdominal symptoms | ICD-9/10 | 7899, 7890, R190 |
| 7 | Diagnoses | Other cancer | ICD-9/10 | 1999, 2080, 2381, C809, 199, 2399, 1991, C772, C7800, C753 |
| 8 | Diagnoses | Respiratory system | ICD-9/10 | 4869, 4939, 7862, 7860 |
| 9 | Diagnoses | Routine encounter | ICD-9/10 | V709, V202, V700 |
| 10 | Diagnoses | Signs and symptoms – ear | ICD-9/10 | 3887, 3801, 3899, 3829 |
| 11 | Diagnoses | Signs and symptoms – Genito urinary system | ICD-9/10 | 5959, 6289, 6049, 6069, 5990, V723, 6202, 6089, N839 |
| 12 | Diagnoses | Signs and symptoms – GI | ICD-9/10 | 5640, 7870, R112, K590, R113, R111 |
| 13 | Diagnoses | Signs and symptoms – hematology | ICD-9/10 | 2889, 2859, D649, D696, D630, D700 |
| 14 | Diagnoses | Signs and symptoms – NOS | ICD-9/10 | 0799, 7865, V725, V720, V729, H471, H498 |

| **Supplementary Table S14.** Eligible encounter categories for carcinomas | | | | |
|---|---|---|---|---|
| **Category number** | **Code type** | **Category name** | **Billing codes** | |
| **Procedure groups** | | | | |
| 1 | Procedure | Anesthesiology | CCI | |
| | | | ACTE | 00255, 41004, 41039, 41045, 41049 |
| 2 | Procedure | Biopsy | CCI | 2MK71LA, 2NQ71BA, 2WY71HA, 2GM71BA, 2FU71HA, 2MD71LA, 2NA71BA, 2NM71BA, 2NF71BA, 2MJ71LA, 2NK71BA, 2MC71LA |
| | | | ACTE | 00187, 00213, 00282, 00849, 09464, 00252, 20117, 00202, 00215, 00515, 00308, 00184, 00237 |
| 3 | Procedure | Cancer-related encounter | CCI | 1AX35HH, 1ZZ35HA, 1LZ35HH |
| | | | ACTE | 00596, 00734, 09168 |
| 4 | Procedure | Cardiovascular related encounter | CCI | 1IS53GR, 1LZ19HH |
| | | | ACTE | 00350, 00765, 08303, 08360 |
| 5 | Procedure | Consult or assessment – other specialty | CCI | |
| | | | ACTE | 00036, 00553, 08936, 09184, 09288, 00030, 00031, 09282, 16105, 16018, 09255, 09281, 09180, 00043, 09250, 09283, 09150, 09165, 09162, 09160, 09170, 15602, 15487, 15486, 15601, 08976 |
| 6 | Procedure | Consult or assessment – pediatrics | CCI | |
| | | | ACTE | 15543, 15165, 15548, 15540, 15550 |
| 7 | Procedure | Critical care | CCI | 1GZ31CA |
| | | | ACTE | 00928, 00912, 00940 |
| 8 | Procedure | CT – abdomen, pelvis | CCI | |
| | | | ACTE | 08263, 08255, 08266, 08703, 08268, 08262 |
| 9 | Procedure | CT – head, neck, spine | CCI | |
| | | | ACTE | 08275, 08258, 08261, 08290, 08259, 08260 |
| 10 | Procedure | Emergency room | CCI | |
| | | | ACTE | 00051, 15212, 15217, 15218, 15355, 15216, 15461, 60702, 09108, 15213, 15210 |
| 11 | Procedure | Exploration and surgical resection – thyroid | CCI | 1FU87NZ, 1FV83NZ, 1FU89NZ |
| | | | ACTE | 06151, 06280, 06284, 06152 |
| 12 | Procedure | Exploration and surgical resection – skin | CCI | 1YT87LA, 1YG87LA, 1YS87LA, 1YV87LA |
| | | | ACTE | 01121, 01221, 01342, 01350, 02367, 01133, 01205, 01132, 01365, 01134 |
| 13 | Procedure | Exploration and surgical resection – other | CCI | 1GK87LA, 1NV89LA, 1MC89LA, 1YM87LA, 2GE70BA, 1FM87VW, 1MC87LA, 1NV89DA, 2OT70DA, 2GM70BA, 1FR87LA |
| | | | ACTE | 00627, 00635, 04161, 20040, 00703, 05043, 05938, 05943, 05947, 03240, 04233, 05937, 05993, 09362, 00324, 05998, 04242, 05995, 09363, 04159, 00697, 05994, 00519, 00691, 04199, 00746, 05201, 05946, 05987, 05988, 05989, 05996, 02083, 05171, 05183 |
| 14 | Procedure | Miscellaneous | CCI | |
| | | | ACTE | 08925, 00356 |
| 15 | Procedure | MRI – head, neck, spine | CCI | |
| | | | ACTE | 08571, 08570 |
| 16 | Procedure | Other imaging | CCI | |
| | | | ACTE | 08601, 08631, 08679, 08439, 08632, 08645, 08692, 08693, 08700 |
| 17 | Procedure | Ultrasound | CCI | |
| | | | ACTE | 08333, 08348, 08358, 08399, 08359, 08383, 08325, 08366, 08335, 08321, 08326, 08330 |
| 18 | Procedure | Xray – head, neck, chest, spine | CCI | |
| | | | ACTE | 08152, 08100, 08125, 08126, 08059, 08128 |
| **Diagnosis groups** | | | | |
| 1 | Diagnoses | Circulatory system | ICD-9/10 | 4299, 7851, 7856 |

| | | | | |
|---|---|---|---|---|
| 2 | Diagnoses | Endocrine and metabolic disorders | ICD-9/10 | 244, 2521, 2468, 2759, 2599, 2754, 7754, 2559, 2789, 2429, 2462, 2500, 2780, 2409, 246, 2423, 2459, 2410, 2419, 2469, 2449, E669, E834, E063, E039, E892, E835, E890, E041 |
| 3 | Diagnoses | Infectious diseases | ICD-9/10 | 7806, R509 |
| 4 | Diagnoses | Mental and neurological diagnoses | ICD-9/10 | F900, 3019, 7804, F411, 346, 7805, 3009, 3139, 3099, 3119, 3469, 3140, 7840, 3000, F900, F419 |
| 5 | Diagnoses | Other cancers | ICD-9/10 | 1999, 1535, 1539, 1749, C809, 2399, 199, 1991, C7800, C7801, C770 |
| 6 | Diagnoses | Other and Malignant melanomas | ICD-9/10 | 1950, 2374, 1420, D34, 172, 1728, C73, 1739, 1619, 1629, 2269, 1729, 2169, 193, 1939, C435, C437, C07, C181, C73 |
| 7 | Diagnoses | Respiratory system | ICD-9/10 | J069, 7841, 7802, 7863, 4781, 4869, 4739, 4909, 4660, 7860, 4939, 7862 |
| 8 | Diagnoses | Routine encounter | ICD-9/10 | V708, Z000, Z008, V202, V709, V700, V909 |
| 9 | Diagnoses | Signs and symptoms – genitourinary system | ICD-9/10 | 7883, 6202, 6262, 7089, 6253, 5990, 5959 |
| 10 | Diagnoses | Signs and symptoms - GI | ICD-9/10 | 5649, 5368, 541, 7870, 5589, 5640, 5409, K590, R113, K37, R111, K353, 540, 5419 |
| 11 | Diagnoses | Signs and symptoms – hematology | ICD-9/10 | 7909, 2859, 2899, 7847, D649 |
| 12 | Diagnoses | Signs and symptoms – NOS | ICD-9/10 | 0799, 6259, 7239, 4741, 7899, 4740, 7231, 7242, 7291, 7245, R104, 7809, 7807, 7865, 0781, 7295, 7890, 4639, J350, K358, 7834, V829, 7199, V729, V725, 7373 |
| 13 | Diagnoses | Signs and symptoms – dermatological | ICD-9/10 | 6868, 6918, 7049, 7822, 0780, 6969, 4481, 6820, 7029, 7099, 7821, 6829, 6929, 8798 |
| 14 | Diagnoses | Signs and symptoms – ears and eyes | ICD-9/10 | 3899, V720, 3820, 3799, 3629, 3723, 3887, 3801, 3810, 3829 |

| **Supplementary Table S15.** Eligible encounter categories for other neoplasms | | | | |
|---|---|---|---|---|
| **Category number** | **Code type** | **Category name** | **Billing codes** | |
| **Procedure groups** | | | | |
| 1 | Procedure | Anesthesiology | CCI | |
| | | | ACTE | 41001 |
| 2 | Procedure | Cancer-related encounter | CCI | 1AX35HH |
| | | | ACTE | |
| 3 | Procedure | Cardiovascular-related encounter | CCI | 1IS53GR, 1IS53LA |
| | | | ACTE | 09308, 08303, 08360 |
| 4 | Procedure | Consult or assessment – other specialty | CCI | |
| | | | ACTE | 09150, 09160, 09170, 09162 |
| 5 | Procedure | Consult or assessment – pediatrics | CCI | |
| | | | ACTE | 15540 |
| 6 | Procedure | Critical care | CCI | |
| | | | ACTE | 00940 |
| 7 | Procedure | CT – chest, abdomen, pelvis | CCI | |
| | | | ACTE | 08263, 08268, 08262 |
| 8 | Procedure | Exploration or surgical resection | CCI | 1GR87QB, 1GV52HA |
| | | | ACTE | 09472 |
| 9 | Procedure | Miscellaneous | CCI | |
| | | | ACTE | 08925 |
| 10 | Procedure | MRI | CCI | |
| | | | ACTE | 08572, 08575, 08570 |
| 11 | Procedure | Other imaging | CCI | |
| | | | ACTE | 08700 |
| 12 | Procedure | Ultrasound | CCI | |
| | | | ACTE | 08331, 08335, 08366, 08321, 08326 |
| **Diagnosis groups** | | | | |
| 1 | Diagnoses | Circulatory system | ICD-9/10 | 4299 |
| 2 | Diagnoses | Infectious diseases | ICD-9/10 | 7806, B370, R509 |
| 3 | Diagnoses | Mental health and neurological diagnoses | ICD-9/10 | F432 |
| 4 | Diagnoses | Other and unspecified malignant neoplasms | ICD-9/10 | 2169, 1709, 1739, 2399, 1719, 199, 1629, 1991, C492 |
| 5 | Diagnoses | Respiratory system | ICD-9/10 | 4869, 5188, 7862, 4660, 4659, 7860, J189, J90, J984 |
| 6 | Diagnoses | Routine encounter | ICD-9/10 | V709, V700 |
| 7 | Diagnoses | Signs and symptoms – GI | ICD-9/10 | R111, R113, K590 |
| 8 | Diagnoses | Signs and symptoms – hematology | ICD-9/10 | D630, D700 |
| 9 | Diagnoses | Signs and symptoms – NOS | ICD-9/10 | 7199, 7599, 7865, 3829, 7890, 5901, V725, V729 |
| 10 | Diagnoses | Signs and symptoms - dermatological | ICD-9/10 | 6929, 7821 |

**Supplementary Table S16.** Encounter category signal strength and lookback window (weeks) derived from control charts for leukemia.

| | Encounter category | Rule 1-4 lookback | Signal strength rule 1-4 | Rule 1-3 lookback | Signal strength rule 1-3 | Rule 1-2 lookback | Signal strength rule 1-2 | Rule 1 lookback | Signal strength rule 1 | Final lookback |
|---|---|---|---|---|---|---|---|---|---|---|
| | **Procedure groups** | | | | | | | | | |
| 1 | Anesthesiology | | | | | 9 | 99.5% | 4 | N/A | 9 |
| 2 | Biopsy | | | | | 14 | 99.4% | 4 | N/A | 14 |
| 3 | Cancer-related encounter | | | 19 | 99.7% | 9 | N/A | 4 | N/A | 19 |
| 4 | Cardiovascular-related encounter | | | 10 | 99.4% | 9 | N/A | 7 | N/A | 10 |
| 5 | Consult or assessment-other speciality | | | 8 | 97.5% | 5 | N/A | 4 | N/A | 8 |
| 6 | Consult or assessment- Paediatrics | 30 | 93.6% | 27 | N/A | 14 | N/A | 12 | N/A | 30 |
| 7 | Critical care | | | | | 5 | 99.4% | 3 | N/A | 5 |
| 8 | CT- chest, abdomen, pelvis | | | | | 5 | 97.5% | 4 | N/A | 5 |
| 9 | CT-head, neck, spine | | | | | 5 | 97.7% | 4 | N/A | 5 |
| 10 | Emergency room | | | 11 | 94.4% | 6 | N/A | 4 | N/A | 11 |
| 11 | Hematology-related encounter | 14 | 99.5% | 12 | N/A | 11 | N/A | 9 | N/A | 14 |
| 12 | Infectious disease encounter | | | | | 7 | 98.9% | 6 | N/A | 7 |
| 13 | Medical genetics | | | | | 11 | 99.8% | 4 | N/A | 11 |
| 14 | Miscellaneous | | | | | 8 | 99.2% | 7 | N/A | 8 |
| 15 | MRI – chest, abdomen, pelvis | | | | | 5 | 92.6% | 4 | N/A | 5 |
| 16 | MRI-head, neck, spine | | | | | 9 | 98.3% | 4 | N/A | 9 |
| 17 | Neurology-related encounter | | | | | 5 | 99.1% | 3 | N/A | 5 |
| 18 | Other imaging | | | | | 17 | 99.4% | 8 | N/A | 17 |
| 19 | Ultrasound – abdomen, pelvis | 12 | 98.3% | 11 | N/A | 10 | N/A | 6 | N/A | 12 |
| 20 | Ultrasound-other | | | | | 17 | 99.2% | 9 | N/A | 17 |
| 21 | Xray-chest, abdomen, pelvis | 16 | 93.0% | | | 8 | N/A | 7 | N/A | 16 |
| 22 | Xray- head, neck, spine | | | | | 10 | 98.1% | 9 | N/A | 10 |
| 23 | Xray- other | | | | | 12 | 90.1% | 6 | N/A | 12 |
| | **Diagnosis groups** | | | | | | | | | |
| 24 | Circulatory system diagnoses | | | 7 | 99.2% | 4 | N/A | 3 | N/A | 7 |
| 25 | Ear-related diagnoses | | | | | 7 | 70.6% | 3 | 81.3% | 3 |
| 26 | Endocrine and metabolic disorders | | | 6 | 99.3% | 4 | N/A | 3 | N/A | 6 |

| | | | | | | | | | | |
|---|---|---|---|---|---|---|---|---|---|---|
| 27 | Infectious disease | 12 | 92.1% | 11 | N/A | 10 | N/A | 8 | N/A | 12 |
| 28 | Leukemia | | | | | 10 | 99.8% | 9 | N/A | 10 |
| 29 | Other cancer | | | | | 9 | 97.6% | 3 | N/A | 9 |
| 30 | Respiratory system | | | | | 8 | 93.9% | 3 | N/A | 8 |
| 31 | Routine examination | | | | | 4 | 83.4% | 3 | N/A | 4 |
| 32 | Signs and symptoms – genitourinary | | | 8 | 93.5% | 6 | N/A | 5 | N/A | 8 |
| 33 | Signs and symptoms – GI | | | 7 | 97.2% | 5 | N/A | 4 | N/A | 7 |
| 34 | Signs and symptoms – hematology | 26 | 98.9% | | | 19 | N/A | 8 | N/A | 26 |
| 35 | Signs and symptoms – musculoskeletal | 17 | 95.2% | 13 | N/A | 11 | N/A | 10 | N/A | 17 |
| 36 | Signs and symptoms – NOS | | | | | 8 | 94.5% | 5 | N/A | 8 |
| 37 | Signs and symptoms – Vision | | | | | 6 | 98.8% | 3 | N/A | 6 |
| 38 | Signs and symptoms – neurology | | | 18 | 92.7% | 4 | N/A | 3 | N/A | 18 |

Note: Gray indicates rule set did not apply; N/A indicates previous rule set met 80% threshold.

**Supplementary Table S17.** Encounter category signal strength and lookback window (weeks) derived from control charts for lymphoma.

| | Encounter category | Rule 1-4 lookback | Signal strength rule 1-4 | Rule 1-3 lookback | Signal strength 1-3 | Rule 1-2 lookback | Signal strength rule 1-2 | Rule 1 lookback | Signal strength rule 1 | Final lookback |
|---|---|---|---|---|---|---|---|---|---|---|
| | **Procedure groups** | | | | | | | | | |
| 1 | Anesthesiology | | | | | 8 | 99.8% | 7 | N/A | 8 |
| 2 | Biopsy – lymph node | | | | | 8 | 98.4% | 3 | N/A | 8 |
| 3 | Biopsy – other | | | | | 10 | 99.7% | 9 | N/A | 10 |
| 4 | Cancer-related encounter | | | | | 12 | 99.4% | 10 | N/A | 12 |
| 5 | Cardiovascular-related encounter | 17 | 98.9% | | | 10 | N/A | 7 | N/A | 17 |
| 6 | Consult or assessment- other specialty | | | | | 10 | 88.3% | 6 | N/A | 10 |
| 7 | Consult or assessment- paediatrics | | | 18 | 92.2% | 10 | N/A | 9 | N/A | 18 |
| 8 | Critical care | | | | | 4 | 96.1% | 3 | N/A | 4 |
| 9 | CT-chest, abdomen, pelvis | | | | | 17 | 99.4% | 11 | N/A | 17 |
| 10 | CT- head, neck, spine | | | | | 12 | 98.9% | 8 | N/A | 12 |
| 11 | Emergency room | | | 12 | 94.8% | 11 | N/A | 7 | N/A | 12 |
| 12 | Medical genetics | | | | | 4 | 97.1% | 3 | N/A | 4 |
| 13 | Miscellaneous | | | | | 8 | 99.0% | 6 | N/A | 8 |
| 14 | MRI- chest, abdomen, pelvis | | | | | 9 | 96.7% | 4 | N/A | 9 |

| | | | | | | | | | | |
|---|---|---|---|---|---|---|---|---|---|---|
| 15 | MRI-head, neck, spine | | | | | 11 | 98.8% | 10 | N/A | 11 |
| 16 | Other imaging | | | | | 17 | 99.9% | 15 | N/A | 17 |
| 17 | Surgical resection and exploration | | | | | 15 | 99.6% | 9 | N/A | 15 |
| 18 | Ultrasound – chest, abdomen, pelvis | | | 29 | 96.3% | 11 | N/A | 10 | N/A | 29 |
| 19 | Ultrasound-other | | | | | 36 | 98.2% | 13 | N/A | 36 |
| 20 | Xray-chest, abdomen, pelvis | | | | | 14 | 97.5% | 7 | N/A | 14 |
| 21 | Xray- head, neck, spine | | | | | 11 | 94.7% | 6 | N/A | 11 |
| 22 | Xray- other | | | | | 12 | 93.8% | 3 | N/A | 12 |
| | **Diagnosis groups** | | | | | | | | | |
| 23 | Circulatory system diagnoses | | | | | 8 | 98.2% | 3 | N/A | 8 |
| 24 | Dermatological conditions | | | 9 | 86.7% | 8 | N/A | 4 | N/A | 9 |
| 25 | Endocrine and metabolic disorders | | | | | 7 | 97.3% | 5 | N/A | 7 |
| 26 | Genito-urinary system | | | | | 10 | 97.3% | 4 | N/A | 10 |
| 27 | Infectious diseases | | | | | 6 | 85.3% | 5 | N/A | 6 |
| 28 | Lymphoma | | | | | 9 | 99.8% | 8 | N/A | 9 |
| 29 | Musculoskeletal system | | | | | 13 | 96.3% | 6 | N/A | 13 |
| 30 | Neurological diagnoses | | | | | 13 | 88.7% | 5 | N/A | 13 |
| 31 | Other cancer | | | | | 14 | 99.4% | 10 | N/A | 14 |
| 32 | Respiratory system | | | | | 14 | 96.4% | 4 | N/A | 14 |
| 33 | Routine examination | | | 15 | 80.2% | 13 | N/A | 5 | N/A | 15 |
| 34 | Signs and symptoms – GI | 11 | 90.9% | | | 7 | N/A | 3 | N/A | 11 |
| 35 | Signs and symptoms – hematology | | | | | 21 | 97.8% | 11 | N/A | 21 |
| 36 | Signs and symptoms – NOS | | | | | 13 | 96.2% | 7 | N/A | 13 |

Note: Gray indicates rule set did not apply; N/A indicates previous rule set met 80% threshold.

**Supplementary Table S18.** Encounter category signal strength and lookback window (weeks) derived from control charts for central nervous system tumours.

| Encounter category | | Rule 1-4 lookback | Signal strength rule 1-4 | Rule 1-3 lookback | Signal strength 1-3 | Rule 1-2 lookback | Signal strength rule 1-2 | Rule 1 lookback | Signal strength rule 1 | Final lookback |
|---|---|---|---|---|---|---|---|---|---|---|
| | **Procedure groups** | | | | | | | | | |
| 1 | Anesthesiology | | | | | 12 | 99.6% | 5 | N/A | 12 |
| 2 | Biopsy | | | | | | | 2 | 98.2% | 2 |
| 3 | Cancer-related encounter | | | | | 10 | 92.2% | 6 | N/A | 10 |
| 4 | Cardiovascular related encounter | | | | | 7 | 97.9% | 6 | N/A | 7 |

| | | | | | | | | | | |
|---|---|---|---|---|---|---|---|---|---|---|
| 5 | Consult or assessment – other specialty | 40 | 80.6% | | | 39 | N/A | 14 | N/A | 40 |
| 6 | Consult or assessment – pediatrics | 45 | 90.0% | 39 | N/A | 11 | N/A | 10 | N/A | 45 |
| 7 | Critical care | | | | | 6 | 98.8% | 5 | N/A | 6 |
| 8 | CT – head, neck, spine | | | | | 30 | 98.9% | 7 | N/A | 30 |
| 9 | Emergency room | | | | | 10 | 96.8% | 9 | N/A | 10 |
| 10 | Medical genetics | | | | | 13 | 94.2% | 2 | N/A | 13 |
| 11 | Miscellaneous | | | | | 5 | 96.6% | 4 | N/A | 5 |
| 12 | MRI – head, neck, spine | | | 21 | 97.4% | 15 | N/A | 7 | N/A | 21 |
| 13 | MRI – other | | | | | 10 | 96.7% | 4 | N/A | 10 |
| 14 | Neurology-related encounter | | | | | 20 | 95.0% | 9 | N/A | 20 |
| 15 | Other brain and CNS procedure | | | | | 6 | 99.9% | 5 | N/A | 6 |
| 16 | Other imaging | | | | | 13 | 99.3% | 5 | N/A | 13 |
| 17 | Respiratory-related encounter | | | | | 10 | 99.1% | 3 | N/A | 10 |
| 18 | Surgical resection – brain and CNS | | | 15 | 97.7% | 14 | N/A | 12 | N/A | 15 |
| 19 | Ultrasound – head, neck | | | | | 6 | 98.4% | 4 | N/A | 6 |
| 20 | Ultrasound – other | 13 | 95.4% | 12 | N/A | 10 | N/A | 6 | N/A | 13 |
| 21 | Vision-related encounter | | | | | 16 | 96.8% | 7 | N/A | 16 |
| 22 | Xray – head, neck, spine | | | | | 18 | 87.0% | 5 | N/A | 18 |
| 23 | Xray – other | | | 10 | 42.0% | 4 | 46.6% | 3 | 50.0% | Not included |
| | **Diagnosis groups** | | | | | | | | | |
| 24 | Brain & CNS tumours | | | | | 10 | 99.2% | 9 | N/A | 10 |
| 25 | Circulatory system | | | 7 | 98.7% | 6 | N/A | 5 | N/A | 7 |
| 26 | Endocrine and metabolic disorders | | | 22 | 93.4% | 10 | N/A | 3 | N/A | 22 |
| 27 | Signs and symptoms – genitourinary system | | | | | 9 | 89.1% | 3 | N/A | 9 |
| 28 | Infectious diseases | | | | | 5 | 92.8% | 4 | N/A | 5 |
| 29 | Neurological disorders | | | | | 11 | 96.0% | 9 | N/A | 11 |
| 30 | Musculoskeletal diagnoses | 12 | 75.7% | | | 8 | 82.0% | 6 | N/A | 8 |
| 31 | Signs and symptoms – nervous system and movement disorders | | | | | 15 | 96.8% | 7 | N/A | 15 |
| 32 | Other cancer | | | 19 | 94.3% | 11 | N/A | 7 | N/A | 19 |
| 33 | Respiratory system | | | | | 6 | 76.9% | 4 | 82.9% | 4 |
| 34 | Routine examination | | | | | 4 | 83.5% | 3 | N/A | 4 |

| | | | | | | | | | | |
|---|---|---|---|---|---|---|---|---|---|---|
| 35 | Signs and symptoms – gastrointestinal tract | | | | | 13 | 92.1% | 8 | N/A | 13 |
| 36 | Signs and symptoms – hematology | | | | | 4 | 96.5% | 3 | N/A | 4 |
| 37 | Signs and symptoms – NOS | 9 | 92.5% | | | 8 | N/A | 6 | N/A | 9 |
| 38 | Signs and symptoms - vision | | | | | 9 | 96.6% | 7 | N/A | 9 |
| 39 | Signs and symptoms – speech, language, hearing | | | | | | | 2 | 83.3% | 2 |
| Note: Gray indicates rule set did not apply; N/A indicates previous rule set met 80% threshold. | | | | | | | | | | |

**Supplementary Table S19.** Encounter category signal strength and lookback window (weeks) derived from control charts for neuroblastoma and peripheral nervous system tumours.

| | Encounter category | Rule 1-4 lookback | Signal strength rule 1-4 | Rule 1-3 lookback | Signal strength 1-3 | Rule 1-2 lookback | Signal strength rule 1-2 | Rule 1 lookback | Signal strength rule 1 | Final lookback |
|---|---|---|---|---|---|---|---|---|---|---|
| | **Procedure groups** | | | | | | | | | |
| 1 | Anesthesiology | | | | | 5 | 99.4% | 4 | N/A | 5 |
| 2 | Biopsy | | | | | 6 | 99.8% | 4 | N/A | 6 |
| 3 | Cancer-related encounter | | | | | 10 | 99.8% | 6 | N/A | 10 |
| 4 | Cardiovascular-related encounter | | | | | 8 | 99.5% | 7 | N/A | 8 |
| 5 | Consult or assessment – other speciality | | | 38 | 96.0% | 13 | N/A | 12 | N/A | 38 |
| 6 | Critical care | | | | | 6 | 98.7% | 4 | N/A | 6 |
| 7 | CT – chest, abdomen, pelvis | | | | | 10 | 99.6% | 9 | N/A | 10 |
| 8 | CT – head, neck, spine | | | | | 8 | 99.0% | 4 | N/A | 8 |
| 9 | Emergency room | | | | | 10 | 96.1% | 6 | N/A | 10 |
| 10 | Miscellaneous | | | | | 8 | 99.4% | 7 | N/A | 8 |
| 11 | Other imaging | | | | | 9 | 99.6% | 8 | N/A | 9 |
| 12 | MRI – chest, abdomen, pelvis | | | | | 8 | 98.1% | 4 | N/A | 8 |
| 13 | MRI – head, neck, spine | | | | | 8 | 98.8% | 7 | N/A | 8 |
| 14 | Consult or assessment - pediatrics | | | | | 15 | 99.1% | 10 | N/A | 15 |
| 15 | Surgical resection of thorax, abdominal, and retroperitoneal structures | | | | | 11 | 98.0% | 6 | N/A | 11 |
| 16 | Ultrasound – chest, abdomen, pelvis | | | | | 40 | 98.7% | 9 | N/A | 40 |
| 17 | Genitourinary-related encounters | | | | | 9 | 99.2% | 5 | N/A | 9 |

| | | | | | | | | | | |
|---|---|---|---|---|---|---|---|---|---|---|
| 18 | Xray – chest, abdomen, pelvis | | | | | 15 | 94.0% | 7 | N/A | 15 |
| | **Diagnosis groups** | | | | | | | | | |
| 19 | Circulatory system | | | | | 8 | 83.6% | 6 | N/A | 8 |
| 20 | Endocrine and metabolic disorders | | | | | 11 | 94.7% | 6 | N/A | 11 |
| 21 | Signs and symptoms – genitourinary system | | | | | 5 | 98.9% | 4 | N/A | 5 |
| 22 | Infectious disease | | | | | 8 | 93.0% | 6 | N/A | 8 |
| 23 | Mental health and neurological diagnoses | | | | | 4 | 98.7% | 3 | N/A | 4 |
| 24 | Musculoskeletal system | | | | | 11 | 97.7% | 7 | N/A | 11 |
| 25 | Neuroblastoma and other PNS tumours | | | | | 7 | 98.2% | 6 | N/A | 7 |
| 26 | Other cancer | | | | | 8 | 99.2% | 6 | N/A | 8 |
| 27 | Respiratory system | | | | | 11 | 95.2% | 3 | N/A | 11 |
| 28 | Routine examination | 6 | 63.5% | | | 4 | 67.0% | 3 | 73.3% | Not included |
| 29 | Signs and symptoms – gastrointestinal tract | | | | | 5 | 94.5% | 3 | N/A | 5 |
| 30 | Signs and symptoms – hematological conditions | | | | | 10 | 91.5% | 4 | N/A | 10 |
| 31 | Signs and symptoms – NOS | | | | | 9 | 98.6% | 7 | N/A | 9 |
| 32 | Signs and symptoms - vision | | | | | 7 | 96.6% | 3 | N/A | 7 |

Note: Gray indicates rule set did not apply; N/A indicates previous rule set met 80% threshold.

**Supplementary Table S20.** Encounter category signal strength and lookback window (weeks) derived from control charts for retinoblastoma.

| | Encounter category | Rule 1-4 lookback | Signal strength rule 1-4 | Rule 1-3 lookback | Signal strength 1-3 | Rule 1-2 lookback | Signal strength rule 1-2 | Rule 1 lookback | Signal strength rule 1 | Final lookback |
|---|---|---|---|---|---|---|---|---|---|---|
| | **Procedure groups** | | | | | | | | | |
| 1 | Biopsy | | | | | | | 2 | 99.8% | 2 |
| 2 | Cancer-related encounter | | | | | 7 | 99.7% | 5 | N/A | 7 |
| 3 | Cardiovascular-related encounter | | | | | 9 | 98.4% | 4 | N/A | 9 |
| 4 | Consult or assessment – other specialty | | | | | 7 | 98.1% | 3 | N/A | 7 |
| 5 | Consult or assessment - pediatrics | | | | | 30 | 86.7% | 5 | N/A | 30 |

| | | | | | | | | | | |
|---|---|---|---|---|---|---|---|---|---|---|
| 6 | CT – head, neck, spine | | | | | 5 | 97.6% | 4 | N/A | 5 |
| 7 | Emergency room | | | | | | | 5 | 95.8% | 5 |
| 8 | Miscellaneous | | | | | | | 2 | 98.9% | 2 |
| 9 | MRI – head | | | | | 8 | 99.4% | 7 | N/A | 8 |
| 10 | Ophthalmology | | | | | 7 | 99.7% | 5 | N/A | 7 |
| 11 | Surgical resection of ocular structures | | | | | 5 | 99.8% | 4 | N/A | 5 |
| 12 | Ultrasound – other | | | | | 9 | 97.9% | 4 | N/A | 9 |
| | **Diagnosis groups** | | | | | | | | | |
| 13 | Infectious disease | | | | | | | 2 | 94.8% | 2 |
| 14 | Mental health and neurological diagnoses | | | | | | | | | Not included |
| 15 | Other cancers | | | | | 4 | 98.8% | 3 | N/A | 4 |
| 16 | Retinoblastoma | | | | | 5 | 99.5% | 4 | N/A | 5 |
| 17 | Routine examination | | | | | 16 | 88.4% | 6 | N/A | 16 |
| 18 | Signs and symptoms – GI | | | | | | | 2 | 98.3% | 2 |
| 19 | Signs and symptoms – NOS | | | | | | | | | Not included |
| 20 | Signs and symptoms – vision | | | | | 6 | 99.1% | 5 | N/A | 6 |

Note: Gray indicates rule set did not apply; N/A indicates previous rule set met 80% threshold.

**Supplementary Table S21.** Encounter category signal strength and lookback window (weeks) derived from control charts for renal tumour.

| | Encounter category | Rule 1-4 lookback | Signal strength rule 1-4 | Rule 1-3 lookback | Signal strength 1-3 | Rule 1-2 lookback | Signal strength rule 1-2 | Rule 1 lookback | Signal strength rule 1 | Final lookback |
|---|---|---|---|---|---|---|---|---|---|---|
| | **Procedure groups** | | | | | | | | | |
| 1 | Anesthesiology | | | | | | | 2 | 99.9% | 2 |
| 2 | Biopsy | | | | | | | 2 | 97.6% | 2 |
| 3 | Biopsy – other | | | | | | | 2 | 99.6% | 2 |
| 4 | Cancer-related encounter | | | | | 16 | 99.7% | 15 | N/A | 16 |
| 5 | Cardiovascular-related encounter | | | | | 14 | 99.0% | 5 | N/A | 14 |
| 6 | Consult or assessment – other specialty | | | | | 40 | 98.3% | 18 | N/A | 40 |

| | | | | | | | | | | |
|---|---|---|---|---|---|---|---|---|---|---|
| 7 | Consult or assessment – pediatrics | | | 11 | 95.3% | 7 | N/A | 6 | N/A | 11 |
| 8 | Critical care | | | | | 7 | 95.2% | 3 | N/A | 7 |
| 9 | CT – chest, abdomen, pelvis | | | | | 17 | 98.7% | 13 | N/A | 17 |
| 10 | Emergency room | | | | | 6 | 99.1% | 5 | N/A | 6 |
| 11 | Genito-urinary-related encounter | | | | | 5 | 93.1% | 4 | N/A | 5 |
| 12 | Miscellaneous | | | | | 5 | 99.4% | 4 | N/A | 5 |
| 13 | MRI – chest, abdomen, pelvis | | | | | 6 | 91.0% | 5 | N/A | 6 |
| 14 | MRI – head, neck, spine | | | | | 5 | 83.5% | 3 | N/A | 5 |
| 15 | Other imaging | | | | | | | | | Not included |
| 16 | Surgical resection of renal tissue | | | | | 5 | 99.2% | 4 | N/A | 5 |
| 17 | Ultrasound | | | 11 | 98.2% | 7 | N/A | 5 | N/A | 11 |
| 18 | Xray – chest, abdomen, pelvis | | | | | 6 | 95.3% | 5 | N/A | 6 |
| | **Diagnosis groups** | | | | | | | | | |
| 19 | Circulatory system | | | | | 6 | 99.3% | 4 | N/A | 6 |
| 20 | Endocrine and metabolic disorders | | | | | | | 2 | 98.9% | 2 |
| 21 | Infectious diseases | | | | | 6 | 98.3% | 5 | N/A | 6 |
| 22 | Other cancer | | | | | 6 | 99.2% | 5 | N/A | 6 |
| 23 | Renal tumours | | | | | 6 | 99.7% | 4 | N/A | 6 |
| 24 | Respiratory system | | | | | | | 2 | 97.8% | 2 |
| 25 | Routine encounters | | | | | 5 | 79.0% | 4 | 83.2% | 4 |
| 26 | Signs and symptoms – Genito-urinary system | | | | | 10 | 99.2% | 8 | N/A | 10 |
| 27 | Signs and symptoms – GI | | | | | 5 | 94.1% | 3 | N/A | 5 |
| 28 | Signs and symptoms – hematology | | | | | 4 | 96.8% | 2 | N/A | 4 |
| 29 | Signs and symptoms – NOS | | | | | 8 | 99.0% | 7 | N/A | 8 |

Note: Gray indicates rule set did not apply; N/A indicates previous rule set met 80% threshold.

**Supplementary Table S22.** Encounter category signal strength and lookback window (weeks) derived from control charts for hepatic tumours

| | Encounter category | Rule 1-4 lookback | Signal strength rule 1-4 | Rule 1-3 lookback | Signal strength 1-3 | Rule 1-2 lookback | Signal strength rule 1-2 | Rule 1 lookback | Signal strength rule 1 | Final lookback |
|---|---|---|---|---|---|---|---|---|---|---|
| | **Procedure groups** | | | | | | | | | |
| 1 | Biopsy | | | | | 6 | 99.0% | 5 | N/A | 6 |
| 2 | Cancer-related encounter | | | | | 5 | 98.8% | 4 | N/A | 5 |
| 3 | Cardiovascular-related encounter | | | | | 33 | 96.5% | 13 | N/A | 33 |
| 4 | Consult or assessment – other specialty | | | | | 7 | 98.5% | 6 | N/A | 7 |
| 5 | Consult or assessment – pediatrics | | | | | 9 | 93.9% | 7 | N/A | 9 |
| 6 | CT | | | | | 6 | 98.6% | 5 | N/A | 6 |
| 7 | MRI | | | | | 6 | 95.5% | 5 | N/A | 6 |
| 8 | Other imaging | | | | | 5 | 97.1% | 4 | N/A | 5 |
| 9 | Miscellaneous | | | | | 5 | 95.8% | 4 | N/A | 5 |
| 10 | Exploration and surgical resection – hepatic tissue | | | | | 4 | 96.1% | 2 | N/A | 4 |
| 11 | Ultrasound – chest, abdomen, pelvis | | | | | 8 | 99.2% | 6 | N/A | 8 |
| 12 | Xray – chest, abdomen, pelvis | | | | | | | | | Not included |
| | **Diagnosis groups** | | | | | | | | | |
| 13 | Circulatory system | | | | | 4 | 97.8% | 3 | N/A | 4 |
| 14 | Endocrine and metabolic disorders | | | | | 4 | 97.9% | 3 | N/A | 4 |
| 15 | Hepatic tumours | | | | | 5 | 99.2% | 4 | N/A | 5 |
| 16 | Infectious disease | | | | | 6 | 96.1% | 4 | N/A | 6 |
| 17 | Other cancer | | | | | 5 | 98.5% | 4 | N/A | 5 |
| 18 | Respiratory system | | | | | 4 | 92.3% | 3 | N/A | 4 |
| 19 | Routine encounter | | | | | 5 | 95.1% | 4 | N/A | 5 |
| 20 | Signs and symptoms – GI | | | | | | | 2 | 92.3% | 2 |
| 21 | Signs and symptoms – hepatology | | | | | 7 | 99.1% | 6 | N/A | 7 |
| 22 | Signs and symptoms – haematology | | | | | | | 2 | 97.9% | 2 |
| 23 | Signs and symptoms – NOS | | | | | 7 | 99.0% | 6 | N/A | 7 |

Note: Gray indicates rule set did not apply; N/A indicates previous rule set met 80%

| Supplementary Table S23. Encounter category signal strength and lookback window (weeks) derived from control charts for bone tumours. | | | | | | | | | | |
|---|---|---|---|---|---|---|---|---|---|---|
| | Encounter category | Rule 1-4 lookback | Signal strength rule 1-4 | Rule 1-3 lookback | Signal strength 1-3 | Rule 1-2 lookback | Signal strength rule 1-2 | Rule 1 lookback | Signal strength rule 1 | Final lookback |
| | **Procedure groups** | | | | | | | | | |
| 1 | Anesthesiology | | | | | 6 | 99.6% | 4 | N/A | 6 |
| 2 | Biopsy | | | | | 5 | 99.7% | 4 | N/A | 5 |
| 3 | Cancer-related encounter | | | | | 22 | 98.0% | 7 | N/A | 22 |
| 4 | Cardiovascular-related encounter | | | 6 | 99.3% | 5 | N/A | 4 | N/A | 6 |
| 5 | Consult or assessment – other specialty | 9 | 98.2% | | | 8 | N/A | 7 | N/A | 9 |
| 6 | Consult or assessment – pediatrics | | | 19 | 94.6% | 7 | N/A | 6 | N/A | 19 |
| 7 | CT – chest, abdomen, pelvis | | | | | 8 | 99.8% | 6 | N/A | 8 |
| 8 | CT – head, neck, spine | | | | | 8 | 97.6% | 4 | N/A | 8 |
| 9 | Emergency room | | | | | 6 | 91.2% | 5 | N/A | 6 |
| 10 | Miscellaneous | | | | | 6 | 98.6% | 5 | N/A | 6 |
| 11 | MRI – chest, abdomen, pelvis | | | | | 6 | 95.2% | 4 | N/A | 6 |
| 12 | MRI – head, neck, spine | | | | | 11 | 98.8% | 3 | N/A | 11 |
| 13 | Other imaging | | | | | 12 | 99.4% | 7 | N/A | 12 |
| 14 | Respiratory-related encounter | | | | | 5 | 94.2% | 4 | N/A | 5 |
| 15 | Surgical exploration and resection | | | | | 5 | 98.3% | 4 | N/A | 5 |
| 16 | Ultrasound | | | | | 18 | 96.7% | 8 | N/A | 18 |
| 17 | Xray – extremities | | | | | 12 | 98.2% | 9 | N/A | 12 |
| 18 | Xray – other | | | | | 11 | 97.7% | 10 | N/A | 11 |
| | **Diagnosis groups** | | | | | | | | | |
| 19 | Bone tumours | | | | | 7 | 98.7% | 6 | N/A | 7 |
| 20 | Endocrine and metabolic disorders | | | | | 4 | 93.5% | 3 | N/A | 4 |
| 21 | Musculoskeletal diagnoses | | | | | 12 | 97.1% | 9 | N/A | 12 |
| 22 | Other cancer | | | | | 7 | 98.3% | 6 | N/A | 7 |
| 23 | Respiratory system | | | | | 4 | 98.2% | 3 | N/A | 4 |
| 24 | Routine encounters | | | | | 10 | 93.9% | 5 | N/A | 10 |

| | | | | | | | | | | |
|---|---|---|---|---|---|---|---|---|---|---|
| 25 | Signs and symptoms – urinary system | | | | | | | 2 | 92.3% | 2 |
| 26 | Signs and symptoms – GI | | | | | | | 2 | 91.9% | 2 |
| 27 | Signs and symptoms – hematology | | | | | 5 | 91.9% | 4 | N/A | 5 |
| 28 | Signs and symptoms – NOS | | | | | 8 | 95.1% | 4 | N/A | 8 |
| Note: Gray indicates rule set did not apply; N/A indicates previous rule set met 80% threshold. | | | | | | | | | | |

**Supplementary Table S24.** Encounter category signal strength and lookback window (weeks) derived from control charts for sarcoma

| | Encounter category | Rule 1-4 lookback | Signal strength rule 1-4 | Rule 1-3 lookback | Signal strength 1-3 | Rule 1-2 lookback | Signal strength rule 1-2 | Rule 1 lookback | Signal strength rule 1 | Final lookback |
|---|---|---|---|---|---|---|---|---|---|---|
| | **Procedure groups** | | | | | | | | | |
| 1 | Anesthesiology | | | | | 6 | 97.6% | 5 | N/A | 6 |
| 2 | Biopsy | | | | | 9 | 99.4% | 8 | N/A | 9 |
| 3 | Cancer-related encounters | | | | | 16 | 99.3% | 15 | N/A | 16 |
| 4 | Cardiovascular-related encounters | | | | | 9 | 99.7% | 7 | N/A | 9 |
| 5 | Consult or assessment – other specialty | | | | | 19 | 96.4% | 9 | N/A | 19 |
| 6 | Consult or assessment – pediatrics | | | 12 | 96.9% | 11 | N/A | 10 | N/A | 12 |
| 7 | Critical care | | | | | 5 | 96.8% | 4 | N/A | 5 |
| 8 | CT – chest, abdomen, pelvis | | | | | 11 | 99.3% | 9 | N/A | 11 |
| 9 | CT – head, neck, spine | | | | | 7 | 99.4% | 6 | N/A | 7 |
| 10 | CT – other | | | | | 8 | 97.0% | 7 | N/A | 8 |
| 11 | Emergency room | | | | | 5 | 96.7% | 3 | N/A | 5 |
| 12 | Exploration and surgical resection – abdomen | | | | | 8 | 98.3% | 4 | N/A | 8 |
| 13 | Exploration and surgical resection – other | | | | | 8 | 98.3% | 6 | N/A | 8 |
| 14 | Genitourinary-related encounter | | | | | 5 | 98.7% | 4 | N/A | 5 |
| 15 | Miscellaneous | | | | | 5 | 98.2% | 4 | N/A | 5 |
| 16 | MRI – abdomen, pelvis | | | | | 6 | 97.4% | 4 | N/A | 6 |
| 17 | MRI – head, neck, spine | | | | | 8 | 98.0% | 7 | N/A | 8 |
| 18 | Other imaging | | | | | 21 | 98.8% | 11 | N/A | 21 |
| 19 | Ultrasound – other | | | | | 24 | 96.9% | 11 | N/A | 24 |
| 20 | Xray – chest, abdomen, pelvis | | | | | | | | | Not included |

| | | | | | | | | | | |
|---|---|---|---|---|---|---|---|---|---|---|
| 21 | Xray – other | | | | | | | | | Not included |
| | **Diagnosis groups** | | | | | | | | | |
| 22 | Circulatory system | | | | | 8 | 93.9% | 4 | N/A | 8 |
| 23 | Endocrine and metabolic disorders | | | | | 4 | 94.4% | 3 | N/A | 4 |
| 24 | Infectious diseases | | | | | 5 | 91.8% | 4 | N/A | 5 |
| 25 | Musculoskeletal diagnoses | | | | | 14 | 97.7% | 10 | N/A | 14 |
| 26 | Other cancer | | | | | 11 | 99.3% | 7 | N/A | 11 |
| 27 | Respiratory system | | | | | 9 | 95.6% | 3 | N/A | 9 |
| 28 | Routine examination | | | 12 | 70.1% | 4 | 84.6% | 1 | N/A | 4 |
| 29 | Signs and symptoms – ear | | | | | | | | | Not included |
| 30 | Signs and symptoms – genitourinary system | | | | | 6 | 96.4% | 4 | N/A | 6 |
| 31 | signs and symptoms – GI | | | | | 6 | 90.4% | 4 | N/A | 6 |
| 32 | Signs and symptoms – haematology | | | | | 8 | 95.1% | 7 | N/A | 8 |
| 33 | Signs and symptoms – NOS | | | | | 12 | 96.9% | 10 | N/A | 12 |
| 34 | Soft tissue and other extraosseous sarcomas | | | | | 8 | 98.4% | 6 | N/A | 8 |

Note: Gray indicates rule set did not apply; N/A indicates previous rule set met 80% threshold.

**Supplementary Table S25.** Encounter category signal strength and lookback window (weeks) derived from control charts for germ cell tumours

| | Encounter category | Rule 1-4 lookback | Signal strength rule 1-4 | Rule 1-3 lookback | Signal strength 1-3 | Rule 1-2 lookback | Signal strength rule 1-2 | Rule 1 lookback | Signal strength rule 1 | Final lookback |
|---|---|---|---|---|---|---|---|---|---|---|
| | **Procedure groups** | | | | | | | | | |
| 1 | Anesthesiology | | | | | 5 | 99.2% | 4 | N/A | 5 |
| 2 | Biopsy | | | | | 4 | 99.2% | 3 | N/A | 4 |
| 3 | Cancer-related encounter | | | | | 7 | 99.4% | 6 | N/A | 7 |
| 4 | Cardiovascular-related encounter | | | | | 6 | 99.2% | 4 | N/A | 6 |
| 5 | Consult or assessment – other specialty | | | | | 21 | 98.6% | 15 | N/A | 21 |
| 6 | Consult or assessment – pediatrics | | | 9 | 98.0% | 8 | N/A | 7 | N/A | 9 |

| | | | | | | | | | | |
|---|---|---|---|---|---|---|---|---|---|---|
| 7 | CT – head, neck, spine | | | | | 8 | 99.5% | 4 | N/A | 8 |
| 8 | CT – chest, abdomen, pelvis | | | | | 7 | 99.7% | 6 | N/A | 7 |
| 9 | Emergency room | | | | | 8 | 97.4% | 7 | N/A | 8 |
| 10 | Exploration and surgical resection | | | | | 4 | 99.7% | 3 | N/A | 4 |
| 11 | Exploration and surgical resection – sex organs | | | | | | | 2 | 99.9% | 2 |
| 12 | Miscellaneous | | | | | 6 | 99.0% | 4 | N/A | 6 |
| 13 | MRI – abdomen, pelvis | | | | | 8 | 99.6% | 7 | N/A | 8 |
| 14 | Other imaging | | | | | 7 | 99.3% | 6 | N/A | 7 |
| 15 | Ultrasound | | | | | 13 | 98.2% | 8 | N/A | 13 |
| 16 | Xray | | | | | 12 | 97.8% | 6 | N/A | 12 |
| | **Diagnosis groups** | | | | | | | | | |
| 17 | Endocrine and metabolic disorders | | | | | 5 | 97.8% | 4 | N/A | 5 |
| 18 | Germ cell tumours | | | | | 7 | 99.7% | 6 | N/A | 7 |
| 19 | Infectious diseases | | | | | | | 2 | 95.3% | 2 |
| 20 | Musculoskeletal diagnoses | | | | | 6 | 90.1% | 4 | N/A | 6 |
| 21 | Mental health and neurological diagnoses | | | | | 7 | 98.8% | 6 | N/A | 7 |
| 22 | Other abdominal symptoms | | | | | 12 | 99.1% | 7 | N/A | 12 |
| 23 | Other cancer | | | | | 8 | 98.2% | 2 | N/A | 8 |
| 24 | Respiratory system | | | | | 4 | 90.4% | 3 | N/A | 4 |
| 25 | Routine encounter | | | | | 8 | 78.6% | 2 | 86.1% | 2 |
| 26 | Signs and symptoms – ear | | | | | | | | | Not included |
| 27 | Signs and symptoms – Genito urinary system | | | | | 11 | 98.0% | 6 | N/A | 11 |
| 28 | Signs and symptoms – GI | | | | | 5 | 91.0% | 4 | N/A | 5 |
| 29 | Signs and symptoms – hematology | | | | | 4 | 96.8% | 3 | N/A | 4 |
| 30 | Signs and symptoms – NOS | | | | | 6 | 97.3% | 4 | N/A | 6 |

Note: Gray indicates rule set did not apply; N/A indicates previous rule set met 80% threshold.

**Supplementary Table S26.** Encounter category signal strength and lookback window (weeks) derived from control charts for carcinoma

| | Encounter category | Rule 1-4 lookback | Signal strength rule 1-4 | Rule 1-3 lookback | Signal strength 1-3 | Rule 1-2 lookback | Signal strength rule 1-2 | Rule 1 lookback | Signal strength rule 1 | Final lookback |
|---|---|---|---|---|---|---|---|---|---|---|
| | **Procedure groups** | | | | | | | | | |
| 1 | Anesthesiology | | | | | 10 | 94.2% | 6 | N/A | 10 |
| 2 | Biopsy | | | | | 34 | 99.4% | 23 | N/A | 34 |
| 3 | Cancer-related encounter | | | | | 27 | 89.5% | 8 | N/A | 27 |
| 4 | Cardiovascular related encounter | | | | | 15 | 96.7% | 10 | N/A | 15 |
| 5 | Consult or assessment – other specialty | 54 | 81.6% | 43 | N/A | 40 | N/A | 20 | N/A | 54 |
| 6 | Consult or assessment – pediatrics | | | | | 15 | 89.7% | 4 | N/A | 15 |
| 7 | Critical care | | | | | 8 | 88.1% | 3 | N/A | 8 |
| 8 | CT – abdomen, pelvis | | | | | 21 | 99.3% | 16 | N/A | 21 |
| 9 | CT – head, neck, spine | | | | | 19 | 98.6% | 14 | N/A | 19 |
| 10 | Emergency room | | | | | 13 | 98.9% | 12 | N/A | 13 |
| 11 | Exploration and surgical resection – thyroid | | | | | 4 | 99.4% | 2 | N/A | 4 |
| 12 | Exploration and surgical resection – skin | | | | | 6 | 96.8% | 2 | N/A | 6 |
| 13 | Exploration and surgical resection – other | | | | | 29 | 99.0% | 12 | N/A | 29 |
| 14 | Miscellaneous | | | | | 5 | 59.4% | 1 | 67.5% | Not included |
| 15 | MRI – head, neck, spine | | | | | 11 | 94.3% | 6 | N/A | 11 |
| 16 | Other imaging | | | | | 16 | 92.9% | 6 | N/A | 16 |
| 17 | Ultrasound | | | | | 24 | 94.5% | 15 | N/A | 24 |
| 18 | Xray – head, neck, chest, spine | | | 17 | 79.8% | 8 | 85.5% | 4 | N/A | 8 |
| | **Diagnosis groups** | | | | | | | | | |
| 19 | Circulatory system | | | | | 14 | 82.0% | 4 | N/A | 14 |
| 20 | Endocrine and metabolic disorders | | | | | 35 | 95.8% | 14 | N/A | 35 |
| 21 | Infectious diseases | | | | | 5 | 75.9% | 2 | 87.1% | 2 |
| 22 | Mental and neurological diagnoses | | | | | | | 2 | 80.7% | 2 |
| 23 | Other cancers | | | | | 13 | 98.8% | 8 | N/A | 13 |
| 24 | Other and Malignant melanomas | | | 18 | 97.3% | 12 | N/A | 11 | N/A | 18 |
| 25 | Respiratory system | | | 13 | 79.6% | 11 | 82.1% | 3 | N/A | 11 |

| | | | | | | | | | | |
|---|---|---|---|---|---|---|---|---|---|---|
| 26 | Routine encounter | 15 | 65.8% | | | 4 | 72.6% | 2 | 75.3% | Not included |
| 27 | Signs and symptoms – genitourinary system | | | | | 4 | 64.1% | 1 | 73.0% | Not included |
| 28 | Signs and symptoms - GI | | | | | 4 | 98.9% | 3 | N/A | 4 |
| 29 | Signs and symptoms – hematology | | | | | | | 2 | 75.4% | Not included |
| 30 | Signs and symptoms – NOS | 11 | 91.3% | | | 9 | N/A | 4 | N/A | 11 |
| 31 | Signs and symptoms – dermatological | | | 16 | 72.6% | 11 | 78.8% | 4 | 84.6% | 4 |
| 32 | Signs and symptoms – ears and eyes | | | | | | | 2 | 73.0% | Not included |

Note: Gray indicates rule set did not apply; N/A indicates previous rule set met 80% threshold.

**Supplementary Table S27.** Encounter category signal strength and lookback window (weeks) derived from control charts for other malignant neoplasms.

| | Encounter category | Rule 1-4 lookback | Signal strength rule 1-4 | Rule 1-3 lookback | Signal strength 1-3 | Rule 1-2 lookback | Signal strength rule 1-2 | Rule 1 lookback | Signal strength rule 1 | Final lookback |
|---|---|---|---|---|---|---|---|---|---|---|
| | **Procedure groups** | | | | | | | | | |
| 1 | Anesthesiology | 5 | Not calculated* | | | | | | | 5 |
| 2 | Cancer-related encounter | 4 | Not calculated* | | | | | | | 4 |
| 3 | Cardiovascular-related encounter | 5 | Not calculated* | | | | | | | 5 |
| 4 | Consult or assessment – other specialty | 6 | Not calculated* | | | | | | | 6 |
| 5 | Consult or assessment – pediatrics | 7 | Not calculated* | | | | | | | 7 |
| 6 | Critical care | | | | | | | | | Not included |
| 7 | CT – chest, abdomen, pelvis | 4 | Not calculated* | | | | | | | 4 |
| 8 | Exploration or surgical resection | 5 | Not calculated* | | | | | | | 5 |
| 9 | Miscellaneous | 4 | Not calculated* | | | | | | | 4 |
| 10 | MRI | 5 | Not calculated* | | | | | | | 5 |
| 11 | Other imaging | | | | | | | | | Not included |
| 12 | Ultrasound | 5 | Not calculated* | | | | | | | 5 |
| | **Diagnosis groups** | | | | | | | | | |
| 13 | Circulatory system | | | | | | | | | Not included |

| | | | | | | | | | | |
|---|---|---|---|---|---|---|---|---|---|---|
| 14 | Infectious diseases | 2 | Not calculated* | | | | | | | 2 |
| 15 | Mental health and neurological diagnoses | | | | | | | | | Not included |
| 16 | Other and unspecified malignant neoplasms | 5 | Not calculated* | | | | | 4 | N/A | 5 |
| 17 | Respiratory system | 5 | Not calculated* | | | | | 4 | N/A | 5 |
| 18 | Routine encounter | | | | | | | | | Not included |
| 19 | Signs and symptoms – GI | 2 | Not calculated* | | | | | | | 2 |
| 20 | Signs and symptoms – hematology | 2 | Not calculated* | | | | | | | 2 |
| 21 | Signs and symptoms – NOS | | | | | | | | | Not included |
| 22 | Signs and symptoms - dermatological | | | | | | | | | Not included |

* Signal-strength thresholds not applied due to low case counts; lookback window selected based on presence of any signal.
Note: Gray indicates rule set did not apply; N/A indicates previous rule set met 80% threshold.

| **Supplementary Table S28.** Derivation of binary variables used to describe index encounters and entry into the diagnostic pathway. | | |
|---|---|---|
| **Encounter type** | **Description** | **Included codes** |
| Emergency department visit | Any billing code for reporting an encounter in the emergency room occurring on the index date. | Defined as ≥1 ACTE code on the index date. Codes relating to emergency department visits were drawn from the list of encounter categories provided in Supplementary Tables S4–S15. Codes include: 00051, 09108, 09046, 15182, 15210, 15212, 15213, 15215, 15216, 15217, 15218, 15355, 15461, 15478, 15598, 16020, 60702. |
| Imaging | All billing codes reporting imaging (e.g., X-ray, CT, ultrasound, MRI, PET) occurring on the index date. | Defined as ≥1 imaging billing code on the index date. Imaging codes were drawn from the imaging-related categories listed in Supplementary Tables S4–S15 (Cancer-specific encounter categories), specifically categories such as MRI – head, neck, spine, and Ultrasound. Complete code lists are provided in those tables. |
| Symptomatic | Any recorded symptom reported in the index date. | Defined as ≥1 ICD-9/ICD-10 symptom code on the index date. Symptom codes were drawn from the diagnosis encounter categories listed in Supplementary Tables S4–S15. Complete code lists are provided in the referenced tables. |
| Predisposition syndrome | The presence of a code associated with a known cancer predisposition syndrome occurring in children and adolescents. Recorded as a binary variable, with codes occurring before date of diagnosis. | Defined as the presence of ≥1 ICD-9/ICD-10 code related to a known cancer predisposition syndrome. Codes included: 7513, 2377, 77081, 7580, 7581, 7598, 2840, Q431, Q873, Q8501, G4731, Z150, Q90.0-9, Q91.0-7, D821, Q96.0-9, Q98.0-4, Q992, Q85.8, G11.3, Q82.8, Q87.1, Q85.1, G11.3, D82.0. |

| **Supplementary Table S29.** Median and interquartile range (IQR) of the treatment interval and total interval | | | | |
|---|---|---|---|---|
| | Observed treatment interval (days) | | Observed total interval (days) | |
| Characteristic | Median | IQR | Median | IQR |
| Sex | | | | |
| Female | 0 | (0 to 5) | 28 | (8 to 80) |
| Male | 1 | (0 to 6) | 23 | (8 to 70) |
| Age group at index | | | | |
| <1 | 1 | (0 to 7) | 22 | (8 to 66) |
| 1-4 | 2 | (0 to 6) | 17 | (6 to 60) |
| 5-9 | 1 | (0 to 4) | 21 | (7 to 62) |
| 10-14 | 1 | (0 to 6) | 25 | (8 to 71) |
| 15-19 | 0 | (0 to 6) | 42 | (14 to 103) |
| Cancer type | | | | |
| Leukemia | 1 | (0 to 3) | 11 | (4 to 44) |
| Lymphoma | 2 | (0 to 8) | 32 | (13 to 65) |
| CNS | 0 | (0 to 4) | 30 | (9 to 119) |
| Neuroblastoma | 4 | (0 to 8) | 20 | (9 to 48) |
| Retinoblastoma | 3 | (0 to 7) | 12 | (7 to 60) |
| Renal tumour | 0 | (0 to 5) | 17 | (7 to 66) |
| Hepatic tumour | 5 | (0 to 9) | 15 | (9 to 34) |
| Bone | 9 | (4 to 19) | 31 | (18 to 52) |
| Sarcoma | 5 | (0 to 14) | 42 | (16 to 95) |
| Germ cell | 0 | (0 to 2) | 16 | (8 to 31) |
| Carcinoma | 0 | (0 to 0) | 100 | (36 to 191) |
| Other neoplasm | 9 | (0 to 31) | 15 | (9 to 37) |
| Year of diagnosis | | | | |
| 2010-2019 | 1 | (0 to 6) | 30 | (9 to 94) |
| 2020-2022 | 1 | (0 to 5) | 22 | (7 to 76) |
| Material Deprivation Index, Quintile | | | | |
| 1 – least | 0 | (0 to 5) | 30 | (9 to 88) |
| 2 | 1 | (0 to 6) | 29 | (7 to 93) |
| 3 | 1 | (0 to 6) | 28 | (8 to 103) |
| 4 | 1 | (0 to 5) | 29 | (9 to 84) |
| 5 – most | 1 | (0 to 7) | 25 | (8 to 85) |
| Social Deprivation Index, Quintile | | | | |
| 1 – least | 1 | (0 to 6) | 32 | (9 to 97) |
| 2 | 1 | (0 to 6) | 29 | (8 to 87) |
| 3 | 1 | (0 to 6) | 24 | (8 to 77) |
| 4 | 0 | (0 to 6) | 28 | (9 to 93) |
| 5 – most | 1 | (0 to 5) | 26 | (6 to 92) |
| Rurality | | | | |
| Montreal (4,000,000 inhabitants) | 1 | (0 to 6) | 28 | (8 to 91) |
| Other large urban centres (>100,000 inhabitants) | 0 | (0 to 5) | 30 | (7 to 88) |
| Regional areas (10,000-100,000 inhabitants) | 1 | (0 to 6) | 30 | (8 to 99) |
| Small towns/rural (<10,000 inhabitants) | 1 | (0 to 6) | 26 | (8 to 87) |
